# Resident microbes shape the vaginal epithelial glycan landscape

**DOI:** 10.1101/2022.02.23.22271417

**Authors:** Kavita Agarwal, Biswa Choudhury, Lloyd S. Robinson, Jenifer E. Allsworth, Amanda L. Lewis, Warren G. Lewis

**Author notes:** Current Affiliations: VaxNewMo LLC, St. Louis, MO 63110, United States of America. <u>Co-corresponding authors:</u>.

## Abstract

Epithelial cells are covered in carbohydrates. This glycan coat or “glycocalyx” interfaces directly with microbes, providing a protective barrier against potential pathogens. Bacterial vaginosis is a condition associated with adverse health outcomes in which bacteria reside in direct proximity to the vaginal epithelium. Some of these bacteria, including *Gardnerella*, produce glycosyl hydrolase enzymes. However, glycans of the vaginal epithelial surface have not been studied in detail. Here we elucidate key characteristics of the vaginal epithelial glycan landscape and determine the impact of resident microbes on the surface glycocalyx. In BV, the glycocalyx was visibly diminished. We show that the “normal” vaginal epithelium displays sialylated *N*- and *O*-glycans by biochemical and mass spectrometric analysis. In contrast, BV-epithelial cell glycans were strikingly depleted of sialic acids, with underlying galactose residues exposed on the surface. Treatment of cells from BV-negative women with recombinant *Gardnerella* sialidases generated BV-like glycan characteristics. Together these data provide the first evidence that the vaginal epithelial glycocalyx is attacked by hydrolytic enzymes in BV. Given the widespread structural and functional roles of sialoglycans in biology and disease, these findings may point to a shared epithelial pathophysiology underlying the many adverse outcomes associated with BV.

## Introduction

Resident vaginal bacteria play key roles in women’s sexual and reproductive health^1,2^. Abundant *Lactobacillus* in the vagina is associated with lower risks of gynecologic and obstetric health complications^3–6^. In bacterial vaginosis (BV), lactobacilli are few in number, and the vagina is instead colonized by diverse microbes, including *Gardnerella*^7–11^. BV is associated with pregnancy loss^12,13^, preterm birth^14–16^, postsurgical infections^17^, pelvic inflammatory disease^18–20^, and sexually transmitted infections including chlamydia, gonorrhea, and HIV^21–23^. Despite the broad impact of BV on gynecologic, sexual and reproductive health, current treatments are inadequate and recurrences are frequent^24^.

In BV, vaginal bacteria live in close proximity to the epithelium^10,25–27^. Epithelial cells covered in bacteria, known as ‘clue cells’, are a major criterion for clinical diagnosis of BV^28,29^. Fluorescence *in situ* hybridization studies have identified some of the attached bacteria, which includes *Gardnerella*, *Atopobium, Mobiluncus,* and BVAB1-3^10,25,30,31^. Normal shedding of vaginal epithelial cells from the superficial layers into the lumen is heightened in BV^32,33^. The bacteria-studded, ruffled, and damaged appearance of these exfoliated cells in BV^33^ supports the hypothesis that bacterial attack on the epithelium might contribute to the wide-ranging health risks posed by BV.

Mammalian epithelial cells are densely coated with carbohydrate chains (glycans)^34–36^, that are often the first point of contact for microbes at mucosal sites^37^. The cellular glycan coat or glycocalyx can provide physical protection from pathogens^38,39^, mediate host-microbe interactions^40^, and regulate immunity^41–44^. Host glycans can be modified in many human diseases^45,46^, including infectious diseases caused by viruses and bacteria^47^. Prior studies have elucidated glycans in human cervicovaginal <u>fluid</u>^48–53^. However, <u>epithelial</u> glycans in the vagina are poorly defined and their functions (and dysfunctions) therefore remain elusive.

Several lines of evidence suggest that the biology of carbohydrates may be important in the underlying physiology of BV. Levels of carbohydrate-degrading enzymes (glycosidases) in vaginal fluid are higher in women with BV compared to those without BV^52,54–56^. Moreover, vaginal fluids with high concentrations of mucin degrading enzymes were more likely to exhibit low viscosity^57^. One of the most prominent glycoside hydrolases in BV is sialidase, which is not only diagnostic for BV^54,58^ but also associated with adverse pregnancy outcomes (e.g. pregnancy loss, preterm birth, placental infection)^55,59,60^. Sialidases liberate sialic acids from their outermost position on mammalian glycans. Bacteria expressing sialidase activity have been isolated from women with BV^54,61^, suggesting vaginal sialidases are likely of bacterial origin. Prior studies have further shown that sialidases from the common BV bacterium *Gardnerella* can remove sialic acids from mammalian glycoproteins such as immunoglobulins and mucins^61,62^. Consistent with sialic acid removal by sialidase, mucus secretions of women with BV have higher levels of liberated sialic acid^61,63^, and an overall depletion of sialic acids attached to glycans^61^. Together the evidence suggests that vaginal bacteria in BV are capable of degrading glycans.

Here we pursue the hypothesis that the vaginal <u>epithelial glycocalyx</u> is an unappreciated target for bacterial hydrolytic enzymes in BV. We use several complementary methods to elucidate the landscape of glycans on the surfaces of epithelial cells derived from human vaginal specimens. The data provide a blue-print for epithelial *N*- and *O*-glycosylation. In addition, evidence is presented that <u>epithelial</u> glycans sustain hydrolytic damage in BV and that BV-like glycan phenotypes can be emulated by treatment of “normal” cells with recombinant *Gardnerella* sialidases.

## Results

### Sialic acids are depleted from the vaginal glycocalyx in BV

To study the vaginal epithelial glycocalyx, the overall properties of epithelial glycans were first examined using transmission electron microscopy (TEM) using a classical method of ruthenium red and osmium tetroxide staining^64,65^. BV status was determined using Nugent’s scoring method by analysis of Gram-stained slides of vaginal specimens^66^. Epithelial cells were isolated from vaginal swab eluates of women with (Nugent score 7-10) or without BV (Nugent score 0-3) as described in the methods. In a pilot study, epithelial cells from a woman without BV (referred to as “No BV’’ in the figures hereafter) had a fuzzy layer of glycocalyx protruding from the VEC membrane (**Fig. 1A**). However, there was little to no glycocalyx evident on the cell surfaces from one BV specimen (**Fig. 1B**), suggesting that the vaginal glycocalyx may be degraded in BV. These observations propelled us to perform a more detailed characterization of the vaginal glycocalyx and its glycan components.

**Fig. 1:**
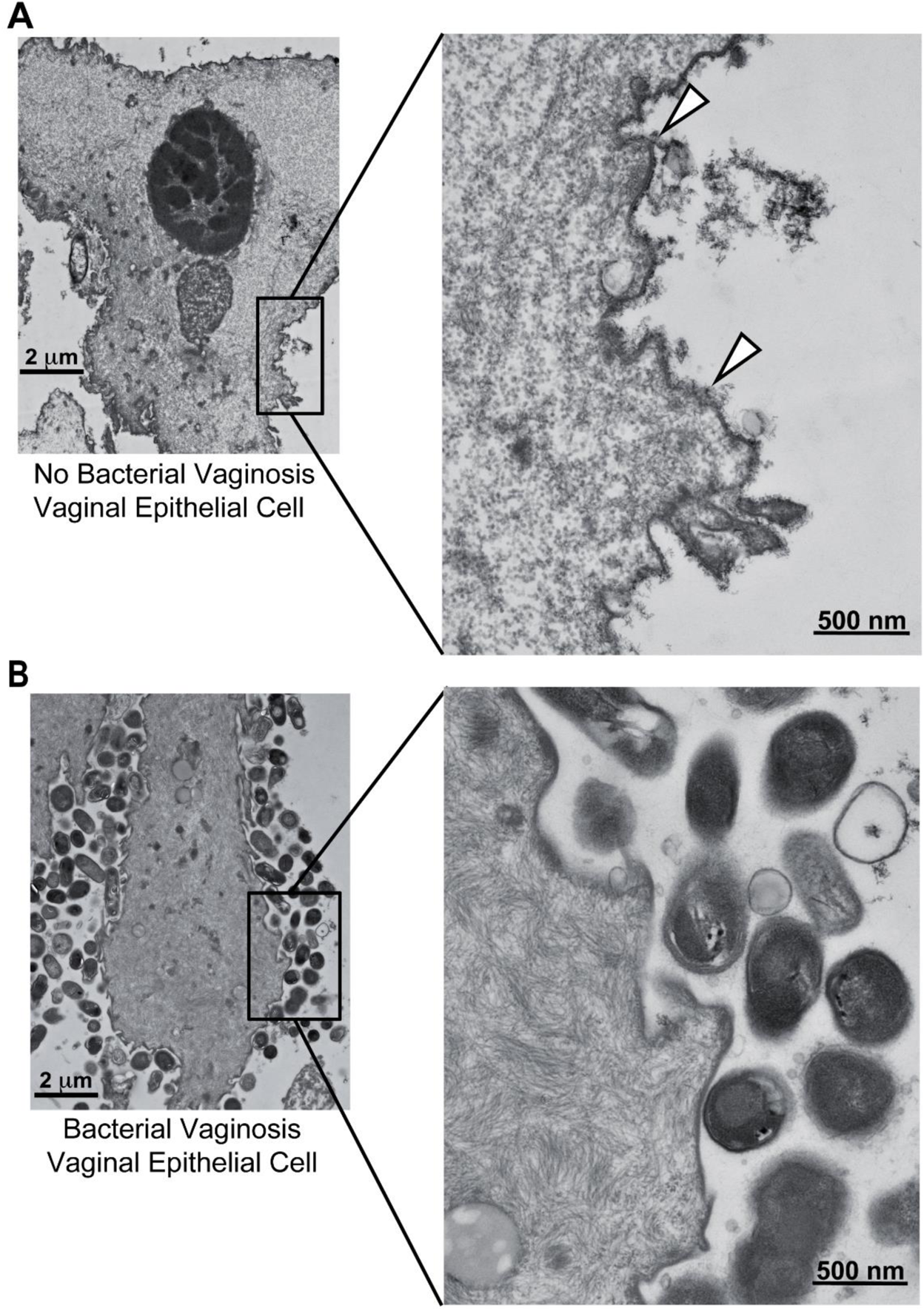
Visualization of the vaginal epithelial glycocalyx in women with and without BV. Representative transmission electron microscopy (TEM) images of vaginal epithelial cell (VEC) stained with ruthenium red and osmium tetroxide. One specimen from (**A**) a woman without BV (Nugent 0-3, referred to as No BV) and (**B**) a woman with BV (Nugent 7-10) were used for staining and microscopy. Arrowheads point to the glycocalyx observed on one VEC from the woman without BV. Data is representative of two independent experiments. Scale bar for images in the left panel is 2 µm and for those in the right panel is 500 nm.

The most abundant sialic acid in humans is *N*-acetylneuraminic acid (Neu5Ac)^42^. Initial investigation of epithelial surface sialylation performed in specimens without BV revealed prominent binding of the *Maackia amurensis* lectin-II (MAL-II) (**Fig. 2A**), which recognizes terminal α2-3-linked sialic acid residues (Neu5Acα2-3Gal)^67^. Specificity was confirmed by pretreating No BV VECs with exogenous sialidase from *Arthrobacter ureafaciens* (*A.u.* sialidase), which eliminated MAL-II binding (**Fig. S1**) and there was minimal background in the secondary alone control (Neutravidin-OG488). Sialidase activity was measured in each vaginal specimen by incubating the eluate with fluorogenic substrate Neu5Ac-4-methyl umbelliferone (4MUSia). In agreement with previous studies^54,56^, we found sialidase levels were significantly higher in samples from women with BV compared to women without BV (**Fig. 2B**). In parallel analyses, eight of nine BV specimens with high sialidase activity (rate of 4MUSia hydrolysis > 0.1), showed negligible MAL-II staining, suggesting extensive depletion of epithelial α2-3-linked sialic acids in BV. In some BV specimens with low sialidase activity, evident MAL-II binding suggested that sialic acid depletion is related to the presence of sialidases in vaginal fluids. The lectin analysis, together with the preliminary electron microscopic analysis, strongly suggest that sialic acids are depleted from the vaginal glycocalyx in women with BV.

**Fig. 2:**
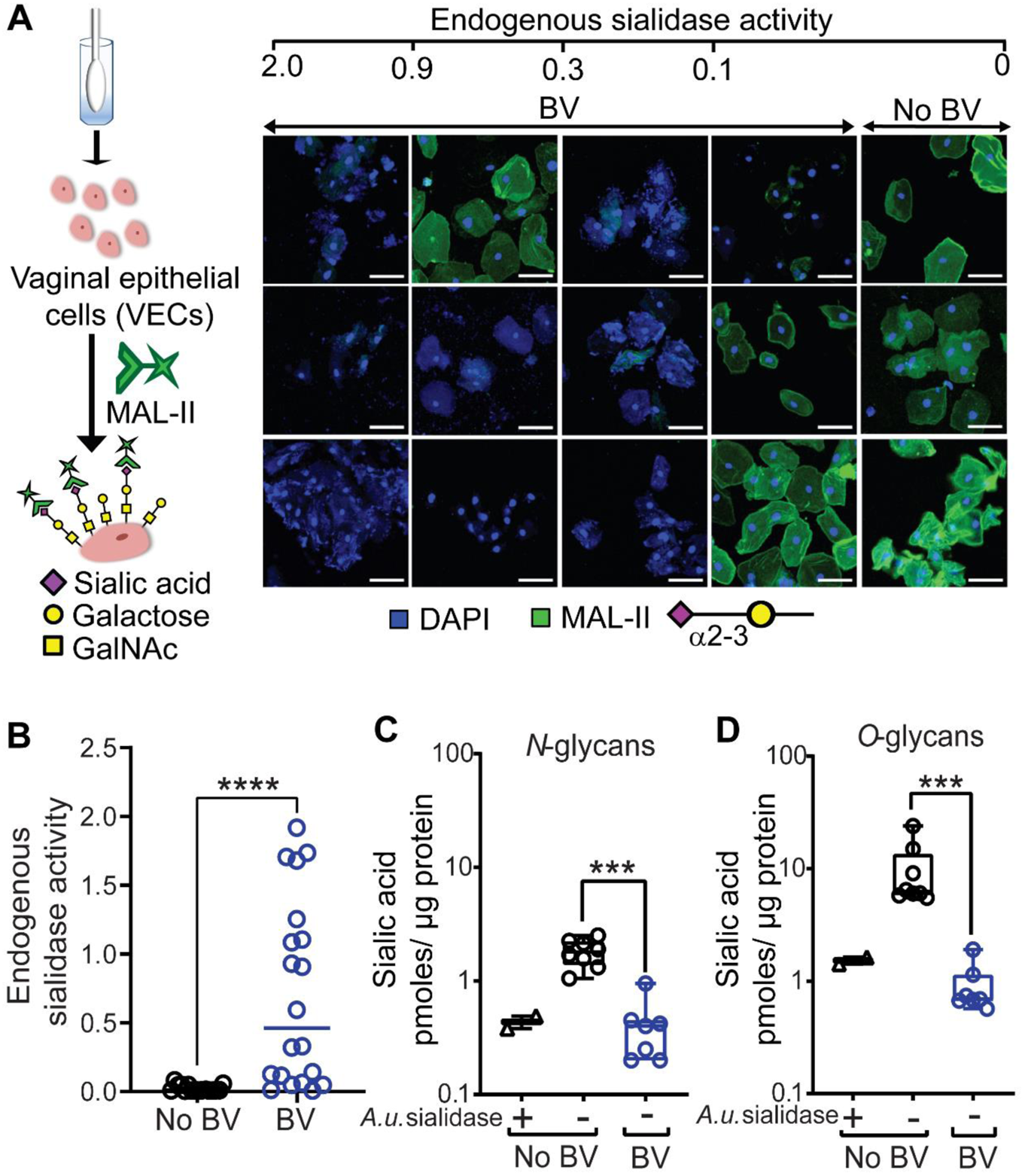
Depletion of sialic acids from vaginal glycans in women with BV. (**A**) Representative confocal images of VECs stained with MAL-II lectin (Green) recognizing α2-3-linked sialic acids from individual women without BV (n=3) and with BV (n=12). Specimens were compared across varying levels of endogenous sialidase activity. Nuclei and bacterial cells (Blue) are stained with DAPI. Scale bars = 50 μm. (**B**) Quantification of endogenous sialidase activity in the vaginal swab eluates using the fluorogenic substrate Neu5Ac-4-methyl umbelliferone (4MU-sialic acid). Data shown is the rate of 4MU hydrolysis and the points represent values for individual women (n=16 without BV and n=20 with BV). Data in **A** and **B** is combined from 3 independent experiments. Images shown in **A** are from a subset of specimens used in **B**. (**C** and **D**) Fluorimetric quantification of 1,2-diamino-4,5-methylenedioxybenzene (DMB)-labelled sialic acid (Neu5Ac) in isolated VEC *N*- and *O*-glycans, using reversed-phase chromatography after mild acid hydrolysis. Graph shows sialic acid quantification on glycans derived from protein extracts of pools of VECs from women with BV (Nugent scores 7-10, n=7 pools from a total N=45 specimens, with 5 or 10 specimens in each pool) and without BV (Nugent scores 0-3, No BV, n=8 pools from a total of N=55 specimens, with 5 or 10 specimens in each pool). Glycans derived from No BV VEC pools pretreated with *A.u.* sialidase were included as a control (n=2 pools with 10 specimens in each pool). Data in **C** and **D** are from same pools of VECs and was combined from 2 independent experiments. Error bars show standard deviation for each group, Mann–Whitney U test was used. ***P < 0.001, ****P<0.0001. *A.u.* sialidase = sialidase from *Arthrobacter ureafaciens.* A total of N=151 specimens were used to generate these data. A subset of VEC pools from **C** and **D** were also used for studies reported in **Fig. 3**, **Fig. 4** and **Table 1**. See methods for pooling rationale.

**Table 1.** Definition of the constituents of *O*-glycans of the vaginal epithelium. We identified the monosaccharide components and linkages of the *O*-glycans of VECs from women without BV by GC-MS analysis of partially methylated alditol acetates (PMAAs). Data is representative of 2 independent experiments. A total of N=10 No BV specimens, combined to form one VEC pool, were used to generate these data. VEC pool generated from these specimens was also used for studies reported in **Fig. 2C, 2D** and to confirm structures of data reported in **Fig. 4**.

| Elution time (min) | Characteristic fragment ions | Assignment<sup>a</sup> | Probable parent O-glycan |
| --- | --- | --- | --- |
| 17.288 | 118,131,162,175 | terminal-Fuc | Fucosylated O-glycans |
| 21.655 | 118, 161, 205 | terminal-Gal | Core-1/Core-2 O-glycan/ T-antigen |
| 24.205 | 190, 161 | 2-Gal | Fucosylated galactose |
| 25.105 | 118,129,161,234 | 3-Gal | Sialylated T-antigen |
| 26.664 | 118, 129,162,189,233 | 6-Gal | Likely Sialylated |
| 27.759 | 130, 246, 290 | 3-GalNAc-itol | Core 1 O-glycan |
| 33.798 | 130, 246, 318 | 3,6-GalNAc-itol | Core 2 O-glycan |
| 35.467 | 117, 159, 233 | 4-GlcNAc | Extended core-2 O-glycan |
| 38.692 | 117, 159, 346 | 3,4-GlcNAc | Lewis antigens |

<sup>a</sup>The respective PMAAs were identified by elution time and their characteristic fragment ions<sup>72</sup>.

The repertoire of glycans on mammalian cell surfaces varies from tissue to tissue and its characterization can be complicated by branching and extensive heterogeneity^68^. Two abundant glycan types modify proteins on mammalian surfaces. *O*-linked glycans are found on serine or threonine residues, whereas *N*-linked glycans are attached to asparagines. To quantitatively assess the degree of sialylation on *N*- and *O*-glycans, epithelial cells were pooled from multiple specimens to yield sufficient material for analysis. Glycans were released from equal amounts of extracted protein for each condition. *N*-glycans were released with PNGase-F under denaturing conditions, while *O*-glycans were released using reductive beta-elimination (see methods). Sialic acids were released from glycans by mild acid hydrolysis, fluorescent derivatized using 1,2-diamino-4,5-methylenedioxybenzene (DMB), and quantified by reverse-phase chromatography by comparison to synthetic standards. Biochemical measurements of sialic acids (Neu5Ac) in these preparations revealed that BV specimens had ~4.5-fold lower sialylation levels on *N*-glycans and ~9-fold lower sialylation levels on *O*-glycans (**Figs. 2C, D**). As a control, VEC pools from women without BV were pretreated with *A.u.* sialidase and washed to remove liberated sialic acid prior to lysate preparation, revealing BV-like *N*- and *O*-glycans with low levels of sialic acid.

### Depletion of negatively charged *N*-linked glycans on epithelial cells in BV

To further characterize epithelial *N*-glycans, we employed anion exchange chromatography to separate neutral and negatively charged glycans. This analysis distinguishes between different types of *N*-glycans [e.g. high mannose (neutral) or mono-, bi-, tri- and tetra-antennary sialylated glycans]. *N*-glycans were isolated from protein extracts of VECs using PNGase-F, labelled with 2-amino benzamide (2-AB) and analyzed by high-performance anion exchange chromatography (HPAEC) with fluorimetric detection (**Fig. 3A**). *N*-glycan standards isolated from the glycoproteins RNaseB and bovine fetuin were analyzed in parallel, illustrating a clear separation between neutral and charged glycans. Neutral *N*-glycan structures were abundant in all women. However, peaks corresponding to negatively charged *N*-glycans were smaller and fewer in samples derived from BV specimens compared to those without the condition (**Fig. 3B**). When the “normal” *N*-glycans were experimentally treated with sialidase, most of the negatively charged (sialylated) peaks disappeared, and the *N*-glycans adopted a BV-like profile (**Fig. 3C**). These results further demonstrate that sialylated epithelial *N*-glycans are found at substantially lower levels in BV. Consistent with analyses presented in **Fig. 2C**, these data further underscore that sialylated *N*-glycans are depleted from vaginal epithelial cells in BV.

**Fig. 3:**
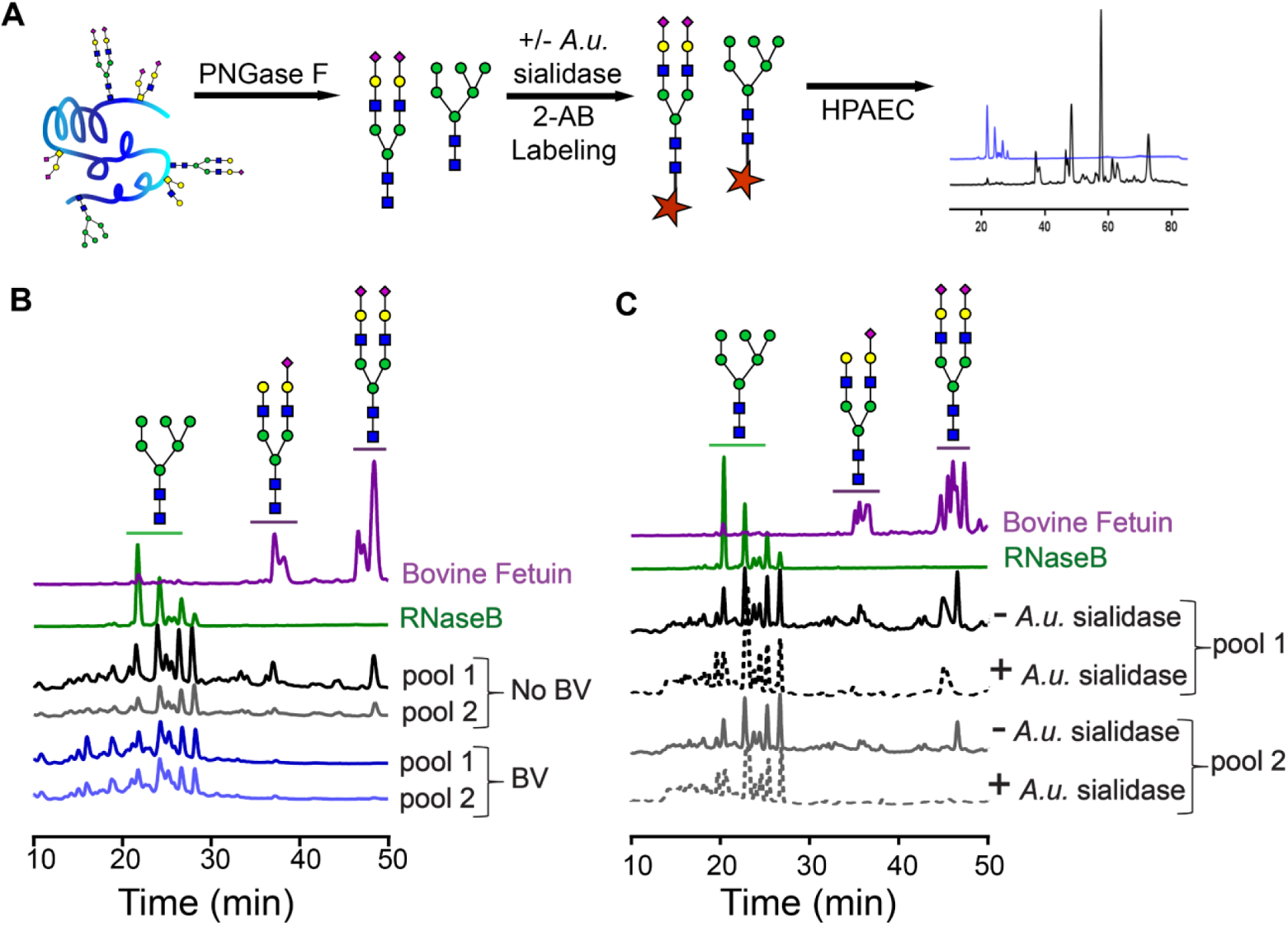
Sialylated vaginal epithelial *N*-linked glycans are depleted in BV. (**A**) Schematic for 2-amino benzamide (2AB) profiling of *N-*linked glycans by high performance anion exchange chromatography (HPAEC). Glycans were released using PNGase-F from protein extracts derived from VECs pools and fluorescently labelled with 2-AB prior to analysis by HPAEC. (**B**) HPAEC profiles of 2-AB labeled *N*-glycans derived from (i) protein standards, with well-known glycan structures, Rnase B and Bovine Fetuin, (ii) 2 pools of No BV VECs (n=10 specimens/pool), (ii) 2 pools of BV VECs (n=10 specimens/pool). (**C**) HPAEC profiles of *N*-glycans derived from (i) Rnase B and Bovine Fetuin, and (ii) pools of No BV VECs (same VEC pools as used in **B**, n=10 specimens/pool) pretreated with commercially available sialidase (+*A.u.* sialidase, dotted line) or with buffer alone (-*A.u*. sialidase, solid line). Structures of different types of *N*-glycans (high mannose, mono- and di-sialylated) are depicted following the NCBI Symbol Nomenclature for Glycans (yellow circle, galactose; green circle, mannose; blue square, *N*-acetylglucosamine; purple diamond, sialic acid). *A.u.* sialidase = sialidase from *Arthrobacter ureafaciens.* A total of N=40 specimens were used to generate these data. VEC pools generated from these specimens were also used for studies reported in **Fig. 2C, 2D** and **Fig. 4**. See methods for pooling rationale.

### *O*-glycans of human vaginal epithelial cells

In women without BV, measurements of sialic acid levels (**Fig. 2D**) suggest that the vaginal glycocalyx contains abundant sialylated *O*-glycans. To identify the relevant *O*-glycan structures of VECs, mass spectrometric (MS) analysis was performed on permethylated epithelial *O*-glycans derived from women without BV.

The most commonly found *O*-glycans in mammals are the mucin type *O*-glycans^69^. Biosynthesis of mucin type *O*-glycosylation is initiated by addition of an alpha-linked *N*-acetylgalactosamine (α-GalNAc) residue to a serine or threonine residue in the peptide backbone^69,70^. Subsequently, the core GalNAc can be extended with other sugars to form several different core structures. Analysis of VEC *O*-glycans using matrix-assisted laser desorption/ionization-time of flight mass spectrometry (MALDI-TOF) revealed that the vaginal glycocalyx contains multiple sialylated *O*-glycan structures. Among these, we observed a small fraction of di-sialylated core 1 (Galβ1-3GalNAc-Ser/Thr, m/z 1256) structures, and relatively high abundance of differentially sialylated and fucosylated core 2 structures (**Fig. 4A**). The core-2 structures are generated by the addition of a β1-6 linked *N*-acetylglucosamine (GlcNAc) to the GalNAc in core 1. Specifically, in “normal” VEC *O*-glycans the highest intensity peak was consistent with a di-sialylated and fucosylated core 2 ion with m/z 1879, containing a potential sialyl Lewis X (sLeX, also known as CD15s) on one antenna. To verify the presence of Lewis X (CD15) structure, we used anti-Lewis X antibody to probe VECs isolated from No BV vaginal specimens with or without prior treatment with exogenous sialidase (*A.u.* sialidase). Analysis by confocal imaging and flow cytometry confirmed the presence of Lewis X structure on the VECs of women without BV (**Fig. S2**). A significant increase in anti-Lewis X binding was observed when VECs were treated with *A.u.* sialidase, suggesting that most of the Lewis X antigens present on “normal” VECs were sialylated, a conclusion also supported by mass spectrometry.

**Fig. 4:**
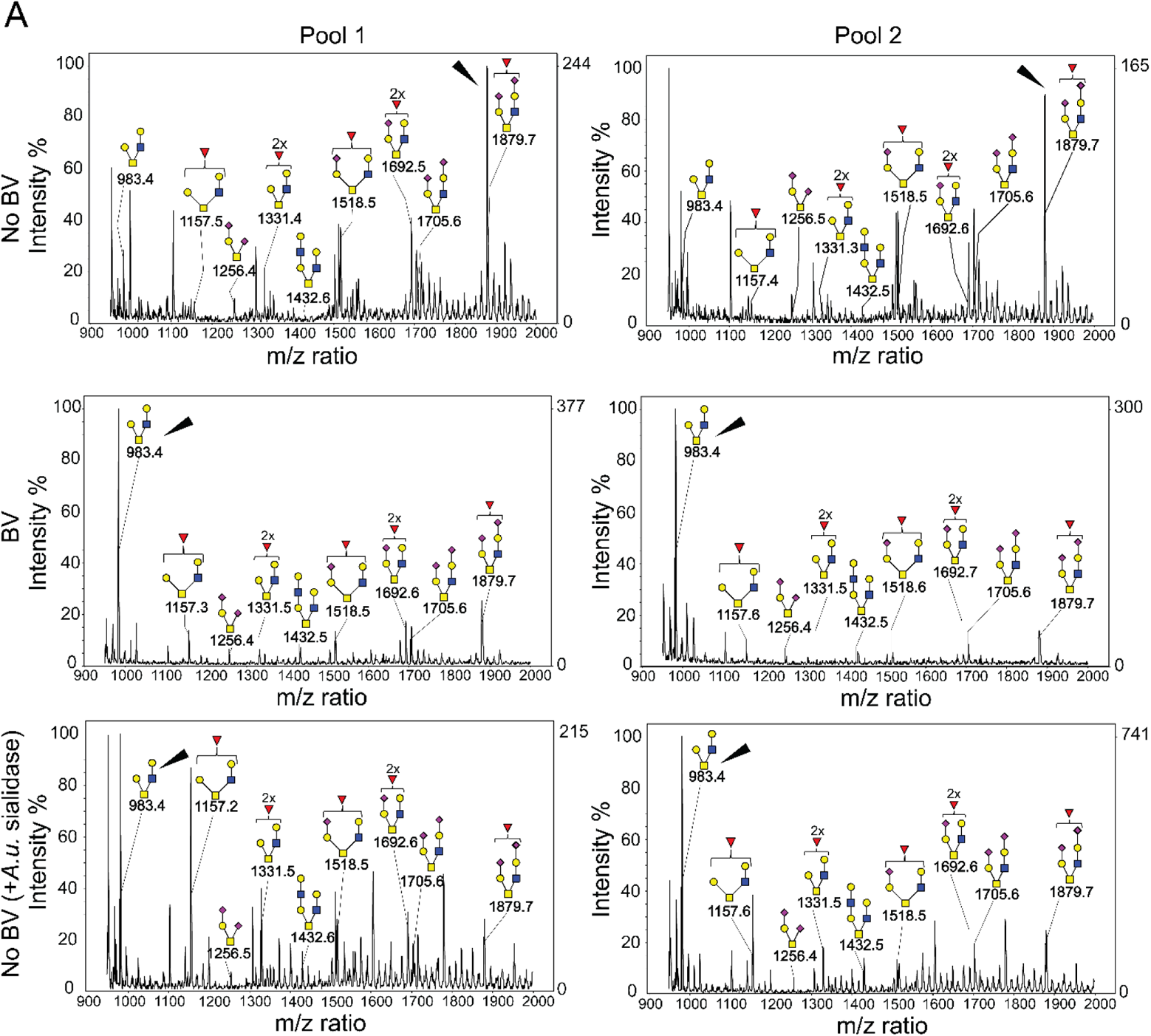
MALDI-TOF spectra of permethylated *O*-glycans present in vaginal epithelial glycocalyx of women with and without BV. (**A**) Mass spectra revealed presence of sialylated core-1 and core-2 glycans in VECs. Asialoglycans dominate in BV (ion m/z = 983), and sialylated glycans dominate under “normal” conditions (ion m/z=1879). Pools of VECs from women without BV pretreated with exogenous sialidase (*A.u.* sialidase) were included as control. Two different pools were analyzed for each condition (n=10 specimens/pool). Structures of different types of *O*-glycans are depicted following the NCBI Symbol Nomenclature for Glycans (red triangle, fucose; yellow circle, galactose; blue square, *N*-acetylglucosamine; yellow square, *N*-acetylgalactosamine; purple diamond, sialic acid). Arrowheads point to assigned *O*-glycan peaks with highest intensity in the spectra. Data is representative of 3 independent experiments. A total of N=60 specimens were used to generate these data. VEC pools generated from these specimens were also used for studies reported in **Fig. 2C, 2D**, **Fig. 3** and **Table 1**. See methods for pooling rationale.

*O*-glycan structures were assigned by tandem MS (MS/MS) analysis of selected precursor ions using a linear ion trap mass spectrometer (LTQ-MS). Fragment ions are described based on the nomenclature proposed by Domon and Costello^71^. The MS/MS fragmentation spectra of permethylated *O*-glycans contained molecular ions characteristic of core 1 and fucosylated core 2 *O*-glycans. We fragmented the sialylated *O*-glycan precursor ion with m/z 1256, yielding a Y-ion corresponding to sialylated T-antigen (Neu5Ac-Galβ1-3GalNAc-Ser/Thr, m/z 881) and a Z-ion corresponding to sialyl-Tn antigen (Neu5Ac2-6GalNAc-Ser/Thr, m/z 659) with much lower intensity (**Fig. S3A**) confirming a core 1 structure. Similarly, fragmentation of the most abundant di-sialylated core 2 ion (m/z 1879) yielded daughter ions with m/z 1504, 1673, 1282 and 1129 confirming a bi-antennary glycan (**Fig. S3A**). Presence of Lewis X on VECs (**Fig. S2**) suggests that the fucose is linked to GlcNAc.

To further investigate *O*-glycan structure, we performed linkage analysis of charged *O*-glycans from VECs confirming the presence of 3-GalNAc-itol, 3,6-GalNAc-itol, and 4-GlcNAc (**Table 1 and Fig. S4**), which is consistent with core 1 and core 2 *O*-glycan structures detected in MS/MS analysis (**Fig. S3**). Detection of both 3-linked Gal and 6-linked Gal is consistent with α2-3, as well as α2-6 sialic acid decoration. Additionally, the presence of terminal fucose and 3,4-GlcNAc supports the presence of Lewis antigen(s) on VECs.

Analysis of *O*-glycans isolated from BV vaginal specimens using MALDI-TOF MS revealed much lower intensity (< 30%) of sialyl Lewis X containing core 2 ion (m/z 1879), as compared to No BV *O*-glycans (**Fig. 4A**). This is consistent with the depletion of sialic acids seen in *O*-glycans in women with BV (**Fig. 2D**). Additionally, corroborating these data we also observed a concurrent higher intensity of a core 2 ion lacking sialic acids (m/z 983) in BV *O*-glycans. In support of this assignment, MS/MS analysis (by LTQ-MS) of the most abundant *O*-glycan ion (m/z 983) yielded daughter ions with m/z 747 (Z-ion), 520 (Y-ion, T-antigen), and 486 (B-ion, LacNAc) confirming the presence of a core 2 structure with LacNAc on one antenna and T-antigen on the other (**Fig. S3A**). Further, MS/MS analysis of the precursor ion with m/z 1157 yielded several fucose containing daughter ions with m/z 921 (most abundant, Fucose{Gal-GlcNAc}), 694 (Fucose-Gal-GalNAc with reducing end) and 660 (Fucose{Gal-GlcNAc}), suggesting the presence of multiple fucosylated glycoforms (**Fig. S3B**). In order to verify if these changes in the molecular ion intensity were due to de-sialylation of glycans, we analyzed *O*-glycans isolated from VECs from women without BV that were pretreated with *A.u.* sialidase (**Fig. 4A**). We found that the MALDI-TOF *O*-glycoprofile of sialidase pretreated VECs was similar to that obtained from women with BV. The highest peak in the *O*-glycans of sialidase pretreated samples corresponded to the desialylated core 2 ion (m/z 983), while the relative intensity of molecular ion m/z 1879 (sialic acid containing core 2 ion) was less than 30%. These data confirm that there is a decrease in abundance of sialylated epithelial *O*-glycans in BV, and suggest a possible role of sialidases in the alteration of vaginal epithelial glycan landscape.

### Exposure of carbohydrates underlying sialic acids on vaginal epithelial cells in BV

Sialic acids are most often present at the terminal position of glycans and can protect the underlying sugars from degradation by glycoside hydrolases^73,74^. In both *N*- and *O*-linked glycans, sialic acids often cap galactose (Gal) residues attached to GlcNAc or GalNAc. To evaluate the impact of BV on such glycans, *if any*, we evaluated terminal Gal residues on the vaginal cell surface using peanut agglutinin (PNA), a lectin that recognizes T antigen (Galβ1-3GalNAc) when the Gal is not capped by sialic acids (**Fig. 5A**)^75^. To distinguish if the PNA epitopes on vaginal epithelial surfaces were masked by sialic acids or simply not present at all, PNA binding was compared between cells from the same sample that were either untreated or pretreated with exogenous sialidase (*A.u.* sialidase). We observed minimal PNA binding to untreated VECs in No BV samples (**Fig. 5B**). However, a significant increase in PNA binding was observed when the VECs from the same sample were treated with sialidase. This suggests that the PNA epitope is present on No BV VECs, but is typically masked by sialic acids. In striking contrast, cells from BV-positive women showed considerable PNA binding even in the absence of sialidase treatment (**Fig. 5B**). Furthermore, PNA binding to VECs from women with BV did not increase upon sialidase treatment, suggesting that the PNA epitopes are mostly not masked by sialic acids in BV. These data corroborate the analytical observation of hypo-sialylated epithelial glycans in BV, and strongly support the conclusion that uncapped Gal residues (as part of the T antigen) are present on epithelial cell surface in BV. Control experiments confirmed PNA selectivity and ruled out non-specific binding (**Fig. S5**). As expected, PNA binding to sialidase-treated epithelial cells from BV-positive women could be out-competed with excess lactose (Galβ1-4-Glc), but not with sucralose. Notably, counter-staining with DAPI revealed that the cells from BV-positive women had large numbers of bacteria covering their surfaces (**Fig. S6**), lending them a pink/purple coloration (**Fig. 5C**). In contrast, the *A.u.* sialidase treated cells from BV-negative women had few adherent bacteria and hence stained red. The BV epithelial cells coated in bacteria are reminiscent of clue cells, a diagnostic feature of BV also observed by wet mount microscopy^28^.

**Fig. 5:**
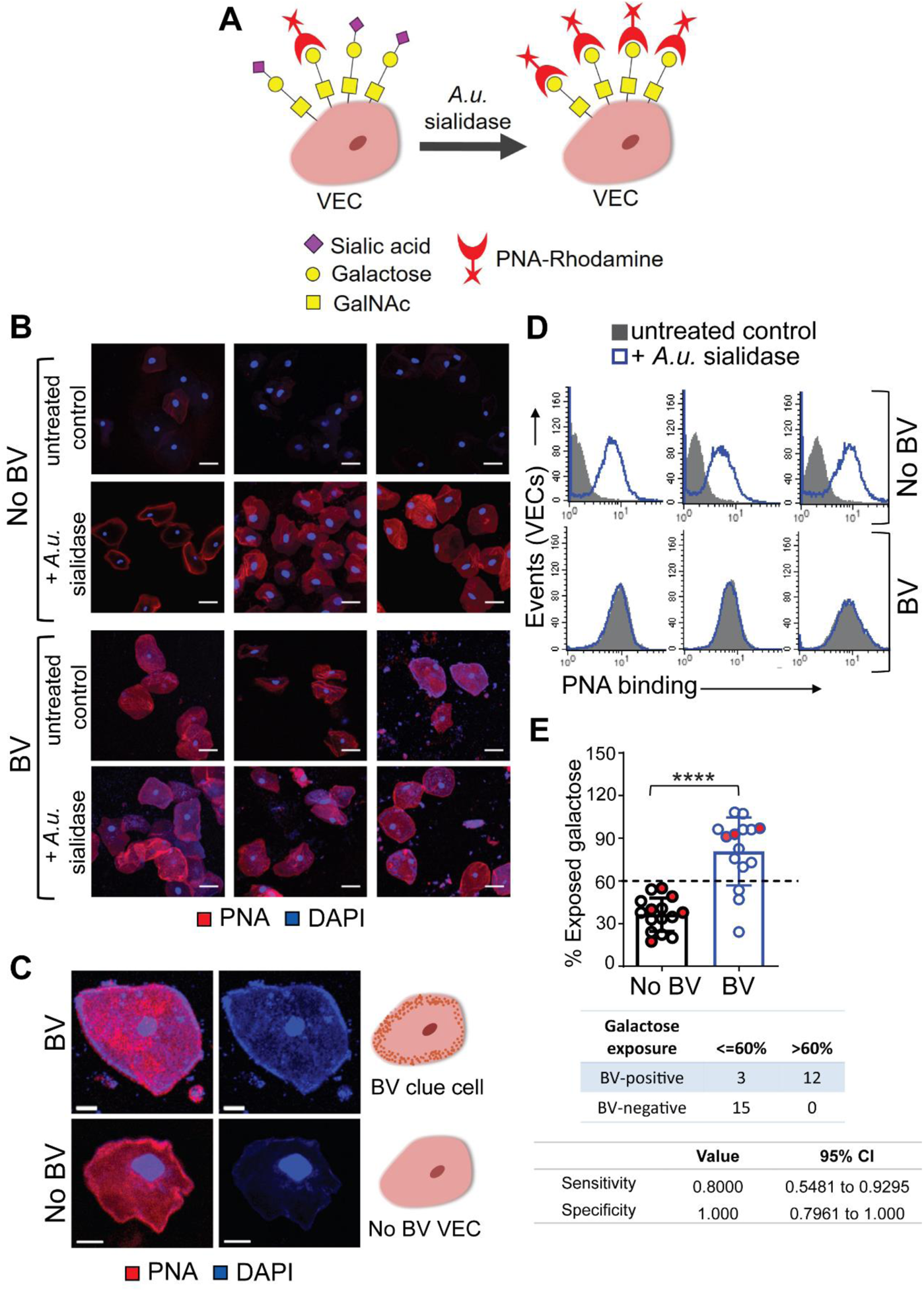
Exposed galactose on the vaginal epithelial cell surface in BV. (**A**) Schematic: PNA-Rhodamine (PNA-Rh) binds to galactose residues, not masked by sialic acids, on VECs treated with or without sialidase. *A.u.* sialidase = sialidase from *Arthrobacter ureafaciens*. (**B**) Representative confocal images of PNA (Red) stained VECs (n=3 individual specimens either with BV or without BV) pretreated with exogenous sialidase (+*A.u.* sialidase) or with buffer alone (*-A.u.* sialidase, untreated control). Nuclei and bacterial cells (Blue) are stained with DAPI. Images shown are representative of multiple fields of view for each specimen. Scale bars = 30 μm. (**C**) Representative image of a BV epithelial cell with surface bacteria observed as blue puncta close to the cell membrane redolent of clue cells. Scale bars = 10 μm. (**D-E**) Flow cytometry quantification of PNA binding to BV and No BV VECs pretreated with exogenous sialidase (*+A.u.* sialidase) or with buffer alone (untreated control). (**D**) Representative histogram overlays are shown from n=6 individual specimens. (**E**) Estimation of percentage cell surface exposed galactose residues, calculated as percentage ratio of mean fluorescence intensity (MFI) of PNA binding to VECs from the same sample in the absence versus presence of *A.u.* sialidase (n=15 samples per group). When possible, data points on the graph represent epithelial cells from individual women (n=22, black/blue circles). However, some data points represent pooled samples from 2-5 women (when recovery yielded insufficient cell quantity for flow cytometry) (n=8 pools, circles filled with red). Data in **A**-**E** was combined from 3 independent experiments. Error bars on the graph show standard deviation for each group. Statistical analysis by Mann–Whitney U, ****P<0.0001. The contingency table below the graph shows sensitivity and specificity values determined using two-sided Fisher’s exact test, with a galactose exposure threshold of 60% compared to the Nugent method. A total of N=46 specimens from individual women were used to generate the data in A-E. See methods for pooling rationale.

**Fig. 6:**
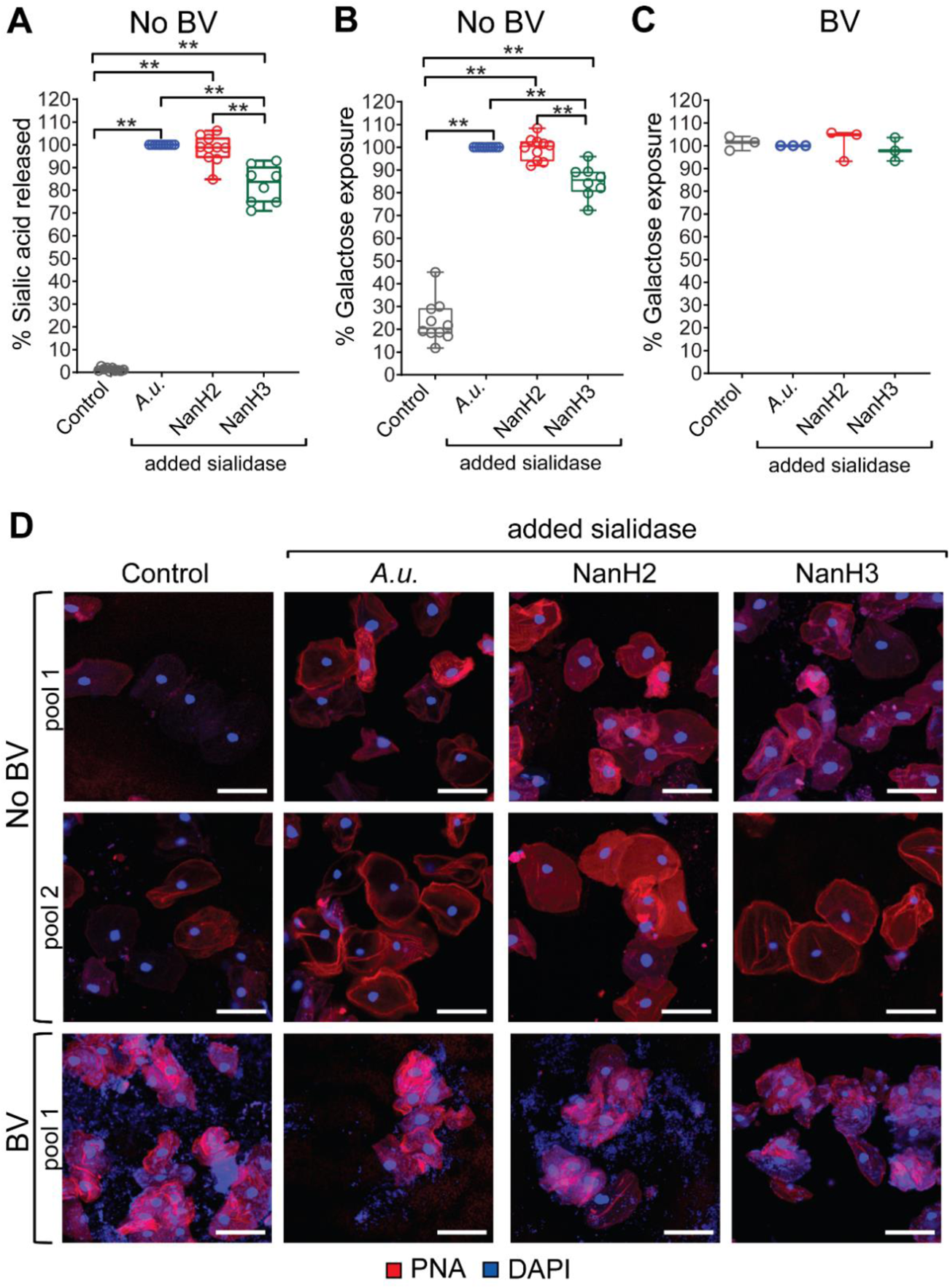
Epithelial cells emulate BV phenotypes when treated with *Gardnerella* sialidases. (**A-D**) Cells from the same pool of VECs, from women with or without BV, were treated with (i) vector control (Control) or (ii) commercial *Arthrobacter ureafaciens* sialidase (*A.u.*) or (iii) recombinant *Gardnerella* NanH2 sialidase (NanH2) or (iv) recombinant *Gardnerella* NanH3 sialidase (NanH3) as indicated in the labels. **(A)** Fluorimetric quantification of sialic acid (Neu5Ac) released from No BV VECs treated with either vector control or sialidase enzymes. (**B, C**) Flow cytometric analysis of PNA binding to VECs. Galactose exposure was assessed by comparing PNA binding to *A.u.* sialidase treated cells from the same pool. (**B**) Galactose exposure on No BV VECs. (**C**) Galactose exposure on BV VECs (n=3 pools, with 2-3 specimens in each pool). **(D)** Representative confocal images of VECs from women with and without BV stained with PNA (Red) lectin under each condition. Nuclei and bacterial cells (Blue) are stained with DAPI. Each row shows analysis of one pooled specimen. Scale bars are 50 µm. Data in **A**-**D** combined from 2 independent experiments. For No BV groups treated with vector control or *A.u.* sialidase or *G.v.* NanH2 sialidase - n=10 pools, with 2 – 5 specimens in each pool; for the No BV group treated with *G.v.* NanH3 sialidase - n=8 pools,, with 2 – 5 specimens in each pool. **p<0.01 (Wilcoxon signed rank test). A total of N=38 specimens from individual women were used to generate the data in **A**-**D**. Data in **7A**, **7B**, and **7D** is from the same No BV VEC pools. Data in **7C** and **7D** is from the same BV VEC pools. See methods for pooling rationale.

To further quantitatively assess the amount of cell surface galactose exposure on epithelial cells from women with and without BV, we measured PNA binding to VECs using flow cytometry. Analysis of the same gated population of cells (**Fig. S7**) from BV and No BV samples showed that PNA binding to cells from BV-positive women was higher compared to the No BV controls in the absence of any exogenous sialidase treatment (**Fig. 5D**). In contrast, epithelial cells from all women were labeled with PNA after sialidase treatment. The extent of exposure of Gal residues in each group was calculated as the percentage ratio of PNA binding to cells in absence versus presence of treatment with *A.u.* sialidase. Using this metric, the differences between BV-negative and BV-positive samples became even more striking. VECs from BV-positive women had a significantly higher (p<0.0001) portion of uncapped Galβ1-3GalNAc (PNA recognized) epitopes compared to cells from BV negative women (**Fig. 5E**).

**Fig. 7:**
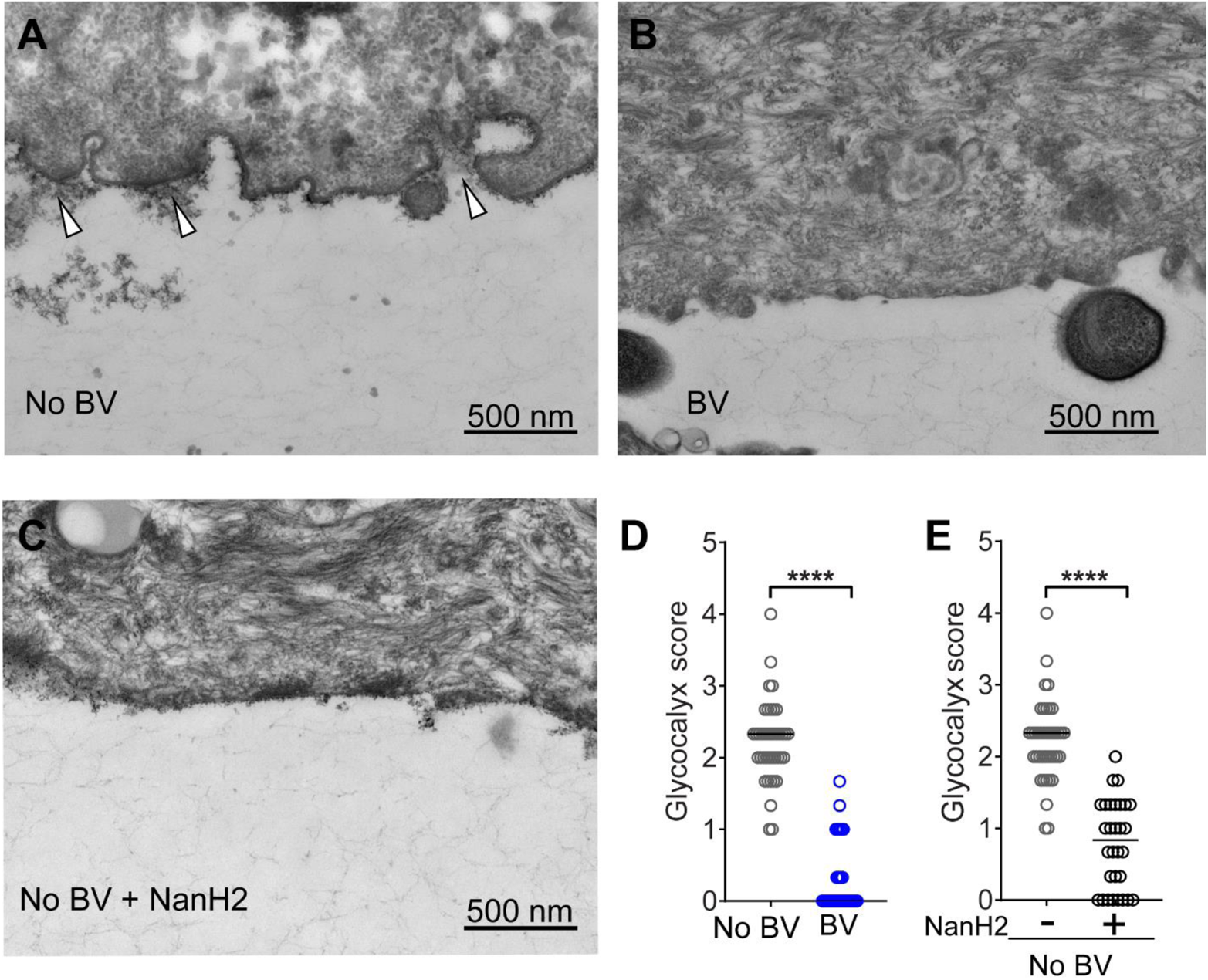
The epithelial glycocalyx is largely absent in BV and *Gardnerella* sialidase diminishes the appearance of the glycocalyx. Representative transmission electron microscopy (TEM) images of VECs from (**A**) No BV specimens - glycocalyx appears as a fuzzy layer close to epithelial cell membrane (indicated with arrowheads), (**B**) BV specimens, and (**C**) No BV specimens - treated with recombinant *Gardnerella* NanH2 sialidase. (**D, E**) Abundance of the glycocalyx was scored on a 0-4 scale, where four is “abundantly present” and zero is “none visible”. A total of ten TEM images were scored from each specimen (scoring rubric in **Fig. S8**). Data points in the graph represent the average of three scores of each image. For (i) No BV; n=40 images from n=4 specimens (same data in shown in **D** and **E**), (ii) BV; n=30 images from n=3 specimens, and (iii) No BV treated with NanH2; n=30 images from n=3 specimens. VECs from three of the No BV specimens were divided in half; one half was treated with recombinant *Gardnerella* NanH2. Images were acquired in a blinded fashion and scored by three observers blinded to the status of the sample. All images were acquired at 25,000 X. Scale bars are 500 nm. ****p<0.0001 (Kruskal-Wallis with Dunn’s multiple comparisons test). A total of N=7 specimens were used to generate these data.

As noted previously^54^, samples from individual BV-positive women also had higher sialidase activity (**Fig. S7**). Correlation analysis, using data from individual un-pooled samples, revealed a direct relationship between the level of sialidase activity and the percentage of exposed galactose on the epithelial cell surface in BV-positive women (**Fig. S7**), analogous to the results presented in **Fig 2A**. Furthermore, in most of the BV samples with high sialidase activity (relative activity >0.5) more than 90% of galactose was found to be exposed in VEC glycans. Interestingly, in a small proportion of BV samples, high galactose exposure (>60%) was also observed on cells from vaginal swabs with very low or no detectable sialidase activity. This suggests that the phenotype of higher PNA binding to VECs may be a more sensitive indicator of BV compared to sialidase, persisting even when sialidase activity (sialidase) is not detectable. In fact, analysis of the sensitivity and specificity of PNA-binding as a potential diagnostic was promising. Based on the distribution of data from samples in both patient groups, we considered 60% galactose exposure as a threshold and compared the data to the Nugent’s method^66^. Using this small sample set, we found that epithelial cell-PNA-binding analysis had a specificity of 100% and sensitivity of 80% (**Fig. 5E**), suggesting this may be a useful area for diagnostic development.

### *Gardnerella* sialidases hydrolyze sialic acids from vaginal epithelial cells and generate BV-like glycan phenotypes

So far we have employed a commercially available recombinant sialidase from *A. ureafaciens* as a control, to demonstrate the potential impact of exogenous sialidase on glycan phenotypes. Next we tested if the more relevant sialidases from *Gardnerella*, often one of the most abundant microbes in BV, could generate BV-like features of glycans.

Specifically, we investigated if *Gardnerella* sialidases degrade glycan components of the epithelial surface, hydrolyzing sialic acids and yielding underlying Galβ1-3GalNAc epitopes recognized by PNA. Briefly, pools of epithelial cells from women without BV were treated with commercial *A.u.* sialidase or recombinant enzymes from *Gardnerella* (NanH2 or NanH3)^62^. In parallel, a proportion of VECs from the same pool were mock treated using material derived from identically prepared *E. coli* containing the empty vector. Sialic acids hydrolyzed from epithelial cells were measured by fluorescent derivatization and HPLC as described earlier for analysis of sialylated *N*- and *O*-glycans. Cells were also subjected to PNA staining. The data show that *Gardnerella* sialidases NanH2 and NanH3 were adept at removing sialic acids from epithelial cells (**Fig. 6A**), revealing PNA-reactive underlying Galβ1-3GalNAc epitopes measured by flow cytometry (**Fig. 6B**). Consistent with findings in **Fig. 5**, BV samples analyzed in parallel had high levels of PNA binding that could not be further increased with treatment by any of the sialidases (**Fig. 6C**). Confocal imaging of PNA-stained cells further underscored the generation of a BV-like phenotype when No BV samples were treated with *Gardnerella* sialidase (**Fig. 6D**). As observed earlier (**Fig. 5C**), cells from BV-positive samples appeared pink/purple in coloration due to adherent bacteria that were stained blue with DAPI, whereas sialidase treated cells from BV-negative samples had red coloration. Overall, these data suggest that sialidases produced by BV-associated bacteria (e.g. *Gardnerella*) result in degradation of vaginal epithelial glycans.

### The glycocalyx is largely absent in BV and is diminished by *Gardnerella* sialidase

Finally, we used classical methods to assess the impact of BV on the vaginal epithelial glycocalyx. VECs were prepared for transmission electron microscopy (TEM) by staining with ruthenium red and osmium tetroxide. Ruthenium red is a polycationic dye that stains acidic polysaccharides and therefore can facilitate visualization of sialic acid containing and other negatively charged glycans on cell surfaces in conjunction with osmium tetroxide, which attaches to vicinal dihydroxylated molecules. Micrographs were collected by a single blinded microscopist and scored by three blinded observers, using the scoring method described in **Fig. S8**, from zero (“none visible”) to four (“highly abundant”). As observed in initial studies (**Fig. 1**), compared to women without BV glycocalyx was diminished in women with BV (**Figs. 7A, B, and Fig. S8**). A striking 95% of the epithelial images from women without BV were scored as having a visible glycocalyx (38/40 score >1). In contrast, epithelial cells of BV specimens were devoid of glycocalyx ~63% of the time (19/30 had a 0 score from all three observers) (**Fig. 7D**).

In parallel with the TEM analysis of glycocalyx in women with and without BV, we also scored blinded images of No BV VECs that had been treated with recombinant purified *Gardnerella* sialidase (NanH2)^62^. We observed that the treatment of “normal” (No BV) VECs with *Gardnerella* sialidase led to a significantly diminished glycocalyx. Similar to what was observed in women with BV (**Figs. 7B, D and Fig. S8**), NanH2 led to a sharp reduction in the appearance of the glycocalyx (**Figs. 7C, E**). Cells treated with *Gardnerella* sialidase had a glycocalyx score less than two in 29/30 (~97%) images (**Fig. 7E**). In, contrast, the untreated group had a score of 2 or greater in 32/40 (~80%) images.

## Discussion

Epithelial surface glycans help maintain homeostasis within mucosal biomes^76–80^. Thus, a full understanding of the physiology of the vaginal epithelium requires detailed knowledge of the structure and functions of glycans presented at the epithelial cell surface. Here we elucidate for the first time the vaginal epithelial glycan landscape using cells isolated directly from human vaginal specimens. Among specimens from women without BV (Nugent score 0-3), epithelial cells had visible glycocalyx in 95% of electron micrographs and analytic approaches revealed prominent sialoglycans on their surfaces. In contrast, cells from women with BV (Nugent score 7-10) displayed epithelial *N*- and *O*-glycans that were strikingly hypo-sialylated and the glycocalyx was absent ~63% of the time. Mass spectrometry showed that epithelial *O*-glycans from BV specimens had lower intensities of ions for sialylated core-1 and core-2 structures, together with higher intensities of desialylated *O*-glycans. Sialylated cell surface *N*-glycans were also depleted in BV, as shown by enzymatic release and chromatographic separation. Staining with multiple glycan-binding lectins in microscopy and flow cytometry confirmed the loss of sialic acids and prominent exposure of terminal galactose on epithelial cell surfaces. Treatment of epithelial cells from women without BV with *Gardnerella* sialidases emulated BV-like phenotypes. Together, the data illustrate that vaginal epithelial cell glycans undergo conspicuous changes in the setting of Nugent-defined BV.

In these experiments, samples were selected based on BV status (Nugent 0-3 and 7-10) from the larger cohort without introducing any strategies to balance the groups for potential confounders. Among the specimens used in this work, women with BV were more likely to be Black, have less education, and a current or past history of sexually transmitted infection (see Table S1). Although not examined here, other studies have found correlates between BV and psychosocial stress^81,82^. We cannot formally rule out contributions by potentially confounding factors to the glycan landscape. However, the concordance between microbes and BV status, enzyme activities, and corresponding degradation of glycan structures strongly argues for a causal relationship between these phenotypes. The finding that *Gardnerella* sialidase acts on epithelial cells to emulate BV glycan phenotypes lends further credence to a causal relationship.

Several lines of evidence strongly suggest that damage to vaginal epithelial glycans is mediated by extrinsic effects of microbiota-derived enzymes in BV. First, sialidase activity in BV specimens corresponds with the presence of sialidase-producing bacterial species, including *Gardnerella*^54,61^. Second, the depletion of α2-3- and α2-6-linked sialic acids from epithelial glycans (shown here) is consistent with the breadth of substrates cleaved by BV sialidases, whether assayed in BV specimens^56^, live *Gardnerella* cultures^61,62^ or using recombinant *Gardnerella* enzymes (NanH2 or NanH3)^62^. Third, BV specimens contain significantly more liberated (free) sialic acid even though there are lower levels of (bound) sialoglycans^61,63^. This is consistent with observations that *in vitro*, strains of *Gardnerella* that express sialidase activity liberate (free) sialic acid into the extracellular space, followed by overall depletion of both free and bound sialic acids^61^. Finally, our data show that recombinant *Gardnerella* sialidases liberate sialic acids from “normal” epithelial cells, resulting in a BV-like epithelial surface (appearance of terminal Gal and visibly diminished glycocalyces). One limitation is that the current experiments do not directly rule out the possibility that host glycosyltransferases or glycosidases might be dysregulated in BV and may make additional contributions to the altered glycosylation status. Nevertheless, together these results strongly suggest a contribution of *Gardnerella*, and potentially other community members, to the dramatic reduction of sialoglycans in BV.

Changes to the glycocalyx could have diverse physiological consequences that may help reveal the pathological mechanisms of BV. From a physical standpoint, the acidic functional groups of sialylated glycans confer a negative charge to the cell surface. Alterations in the organization and net charge contributed by sialic acids are understood to contribute to pathophysiologies of the blood and renal systems^83–87^, and could likewise impair functions of the vaginal epithelium. In addition to physical effects, liberation of cellular sialic acids provides new opportunities for bacterial nutrient acquisition and colonization, including some opportunistic pathogens lacking sialidase activity^61,88–90^. Desialylation may also modify glycan-receptor interactions influencing microbial adhesion, inflammatory responses, or other aspects of host-microbe biology. Viral, bacterial, or eukaryotic pathogens linked with BV might use lectins to exploit the newly accessible glycan epitopes (e.g. exposed Gal residues). For example, *Fusobacterium nucleatum* encodes multiple functionally characterized galactose-recognizing adhesins^91,92^ and colonizes women with BV more often than women without BV^9,14^. The relative absence of sialic acids and exposure of galactose on the BV epithelium could therefore lead to improved colonization by *F. nucleatum* or other potential pathogens linked with BV. Changes in the glycoprotein landscape of the cervicovaginal epithelium may also impact recognition by endogenous lectin receptors of the host. For example, the sialic acid-binding Siglecs and the galactose-binding Galectins are expressed in the female reproductive tract^93–95^ and are known to modulate cellular responses such as inflammatory activation and cell death in other physiological settings^42,96–100^.

These findings may provide new insights into the biology of this prevalent condition, which is little understood despite its widespread impact on sexual, reproductive, and perinatal health. Even though available antibiotics reduce the symptoms of BV, recurrences occur in most women within 12-months of treatment^101^. Future work should investigate if long term efficacy following treatment is contingent upon rehabilitating damaged glycans. If so, the restoration of normal glycosylation might be a valuable test of cure or target for drug development. Additional studies should leverage relevant experimental models and clinical cohorts to understand how specific changes to the glycocalyx may contribute to cellular signaling and function, and how these relate to the biological and clinical facets of BV.

## Supplemental data

**Fig. S1:**
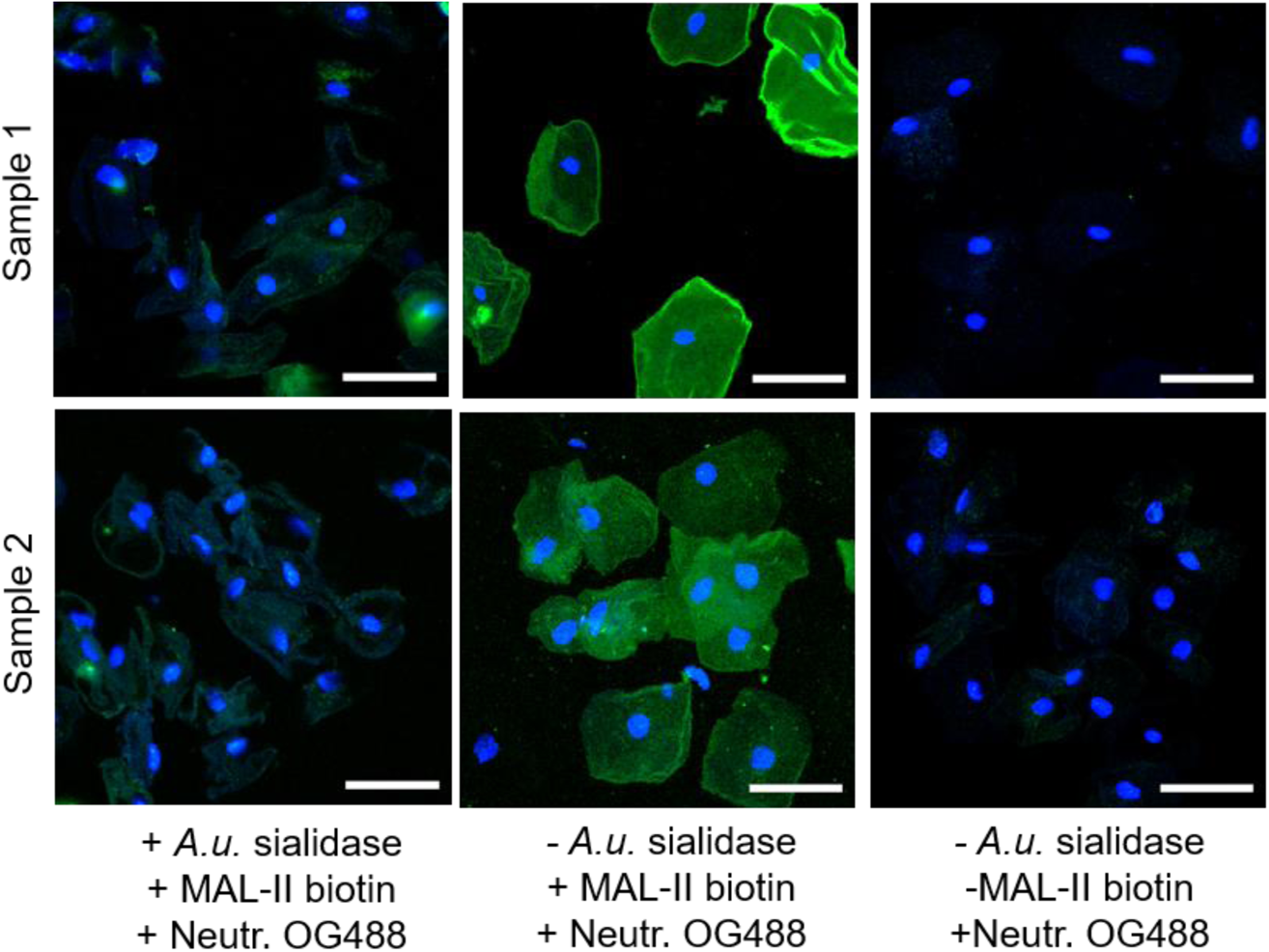
MAL-II binding to VECs from women without BV. Confocal images of VECs from women without BV (Nugent 0-3) that were stained either with biotinylated-MAL-II lectin followed by neutravidin-OG488 (Neutr. OG488) or the neutravidin-OG488 alone (right). VECs pre-treated with exogenous sialidase (*A.u.* sialidase) were included to validate selective binding of MAL-II to sialic acids. Neutravidin-OG488 shows minimal non-specific binding to VECs in the absence of MAL-II lectin. Images shown are representative of multiple fields of view. Scale bars = 50 µm.

**Fig. S2:**
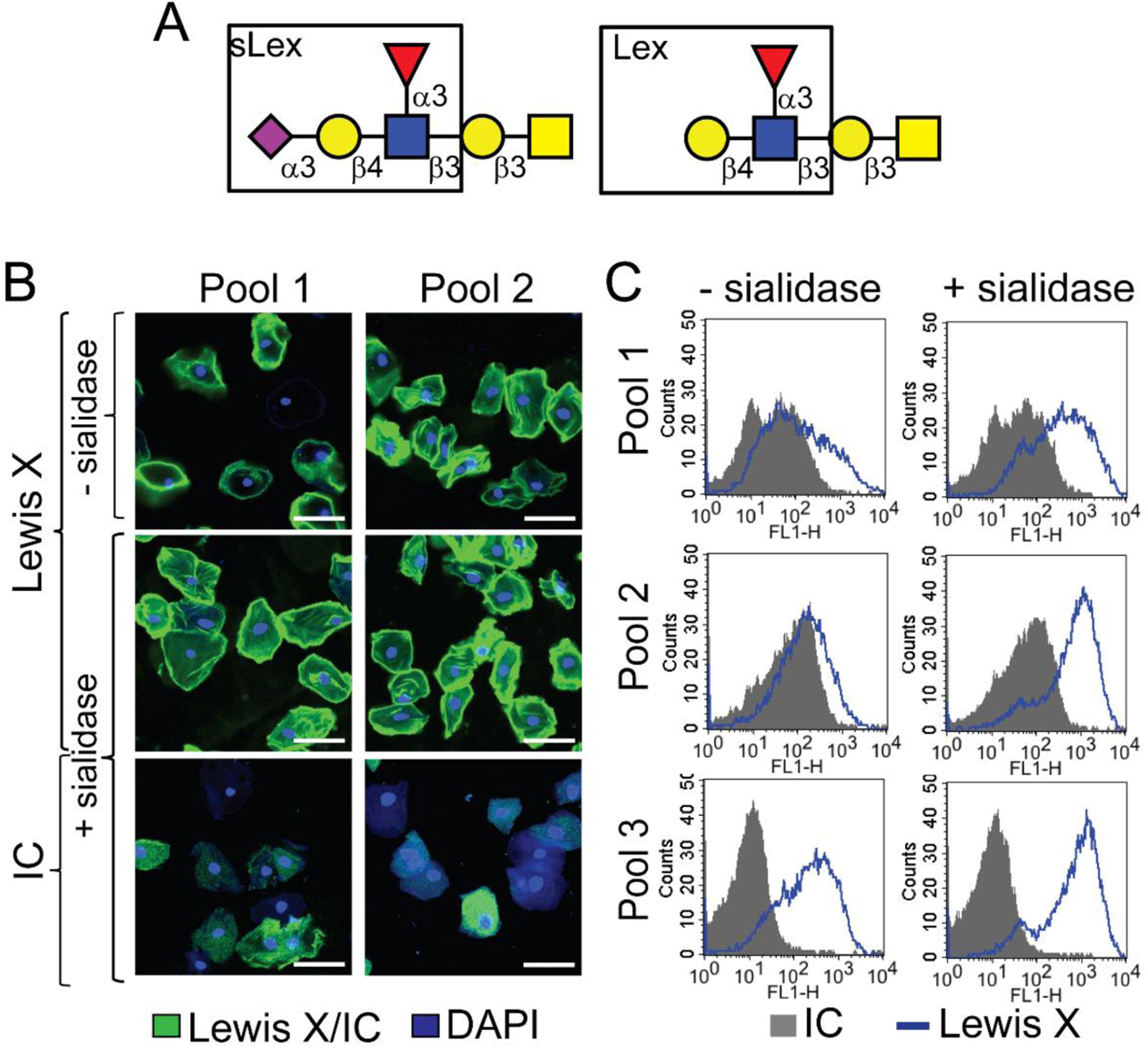
Analysis of Lewis X (CD15) antigen on vaginal epithelial cells. Cells from the same pool of VECs, isolated from vaginal specimens of women without BV (Nugent 0-3), were treated with exogenous sialidase (*A.u.* sialidase) or buffer, followed by staining with anti-Lewis X or isotype control. (**A**) Structures of sialyl Lewis X (sLeX) and Lewis X (LeX) antigens. (**B**) Representative confocal images of two separate pools of VECs. Green = Lewis X or Isotype Control (IC), Blue = DAPI. Scale bars = 50 µm. (**C**) Histograms show binding of the Lewis X antibody to VECs is enhanced upon treatment with exogenous sialidase (*A.u*. sialidase). Grey = IC, Blue = anti-CD15. Data is representative of 3 independent experiments. A total of N=18 specimens from individual women were used to generate the data in **B**-**C**.

**Fig. S3:**
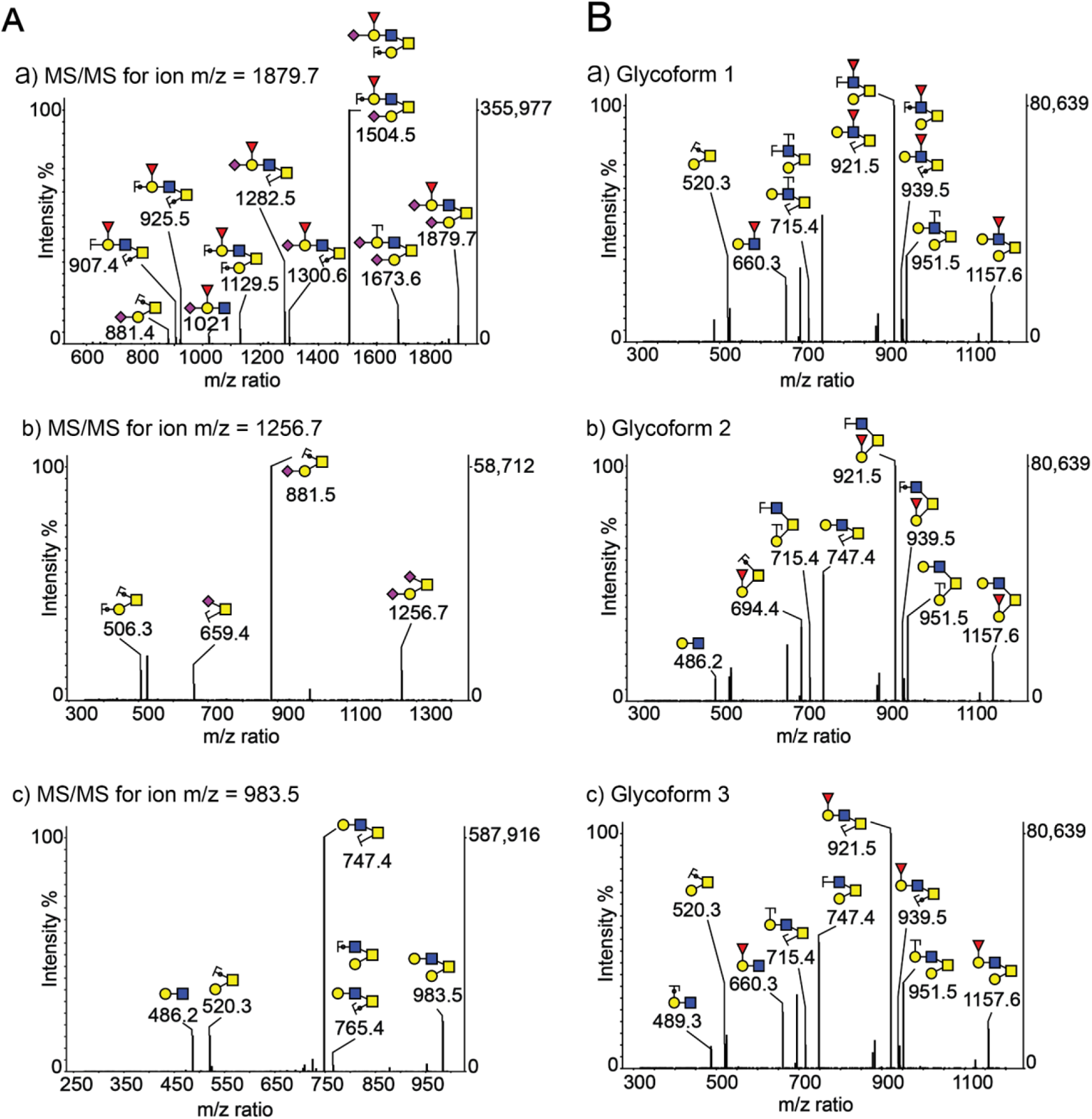
MS/MS spectra of *O*-glycans present in the vaginal epithelial glycocalyx. Representative MS/MS data for permethylated *O*-glycans indicate presence of core-1 and core-2 structures. (**A**) MS/MS for ion m/z ~a) 1878, b) 1257, and c) 983, indicates presence of di-sialylated and fucosylated core-2 *O*-glycan structure, di-sialylated core-1 *O*-glycan structure, and asialo core-2 containing *O*-glycans respectively. (**B**) MS/MS for ion m/z ~1158 indicates presence of three different glycoforms - a, b, and c, with fucose attached either to *N*-acetylglucosamine or galactose residue. Structures are depicted following the NCBI Symbol Nomenclature for Glycans (red triangle, fucose; yellow circle, galactose; blue square, *N*-acetylglucosamine; yellow square, *N*-acetylgalactosamine; purple diamond, sialic acid). The circles superimposing the bonds mean that the oxygen atoms of these glycosidic bonds are included in the fragment, i.e. they are Y-ions or C-ions.

**Fig. S4:**
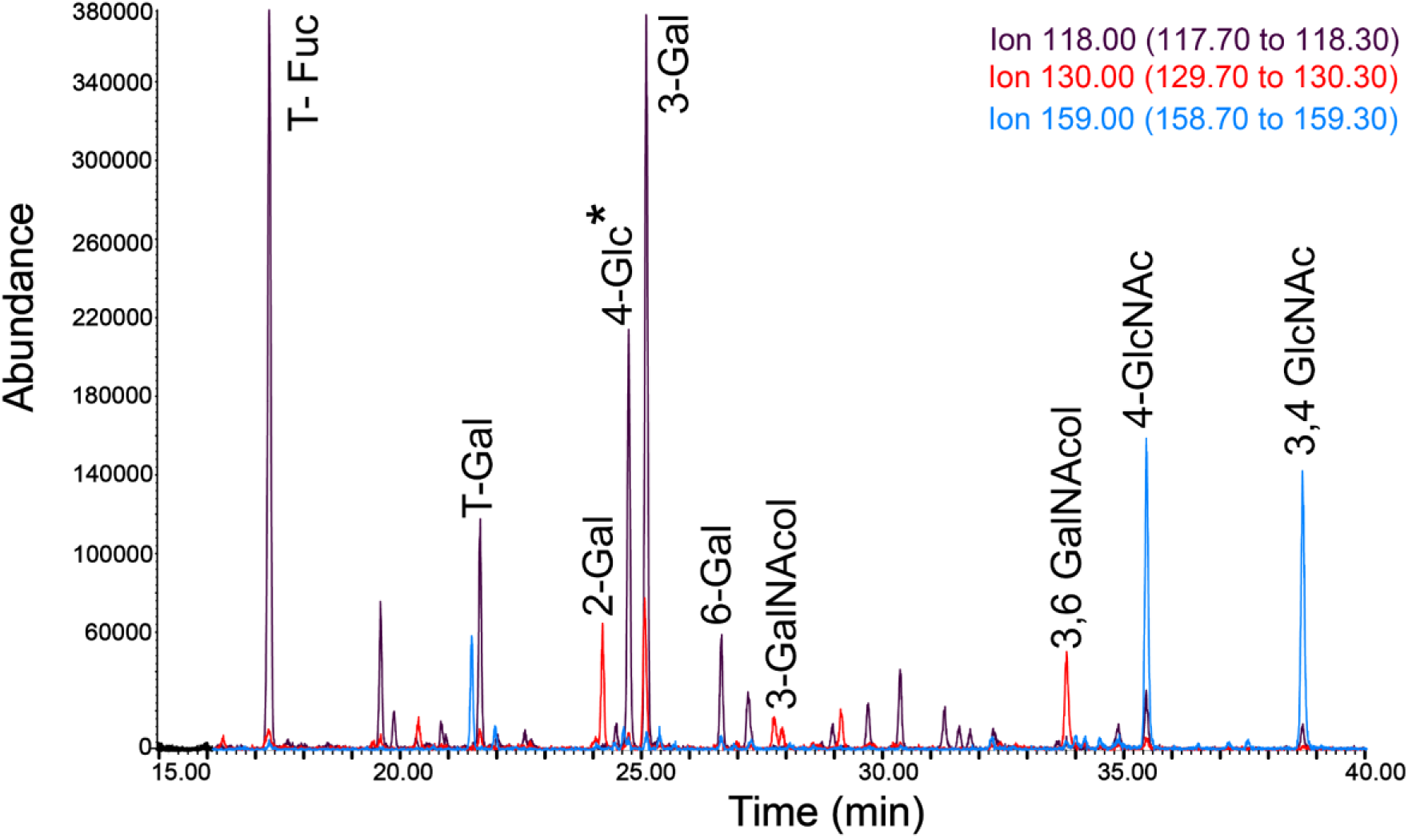
GC-MS linkage analysis confirms the annotation of selected precursor ions from the VEC *O*-glycome. (**A**) Gas chromatogram of charge separated *O*-glycans from a pool of VECs of women with *Lactobacillus* dominated microbiota. Colors indicate ion scans to identify peaks containing monosaccharide fragment ions of a given m/z as indicated in the legend on the upper right corner. Exact elution times for each peak are indicated in Table 1. *4-Glc - indicates a contaminating peak from glycogen present in vaginal specimens.

**Fig. S5:**
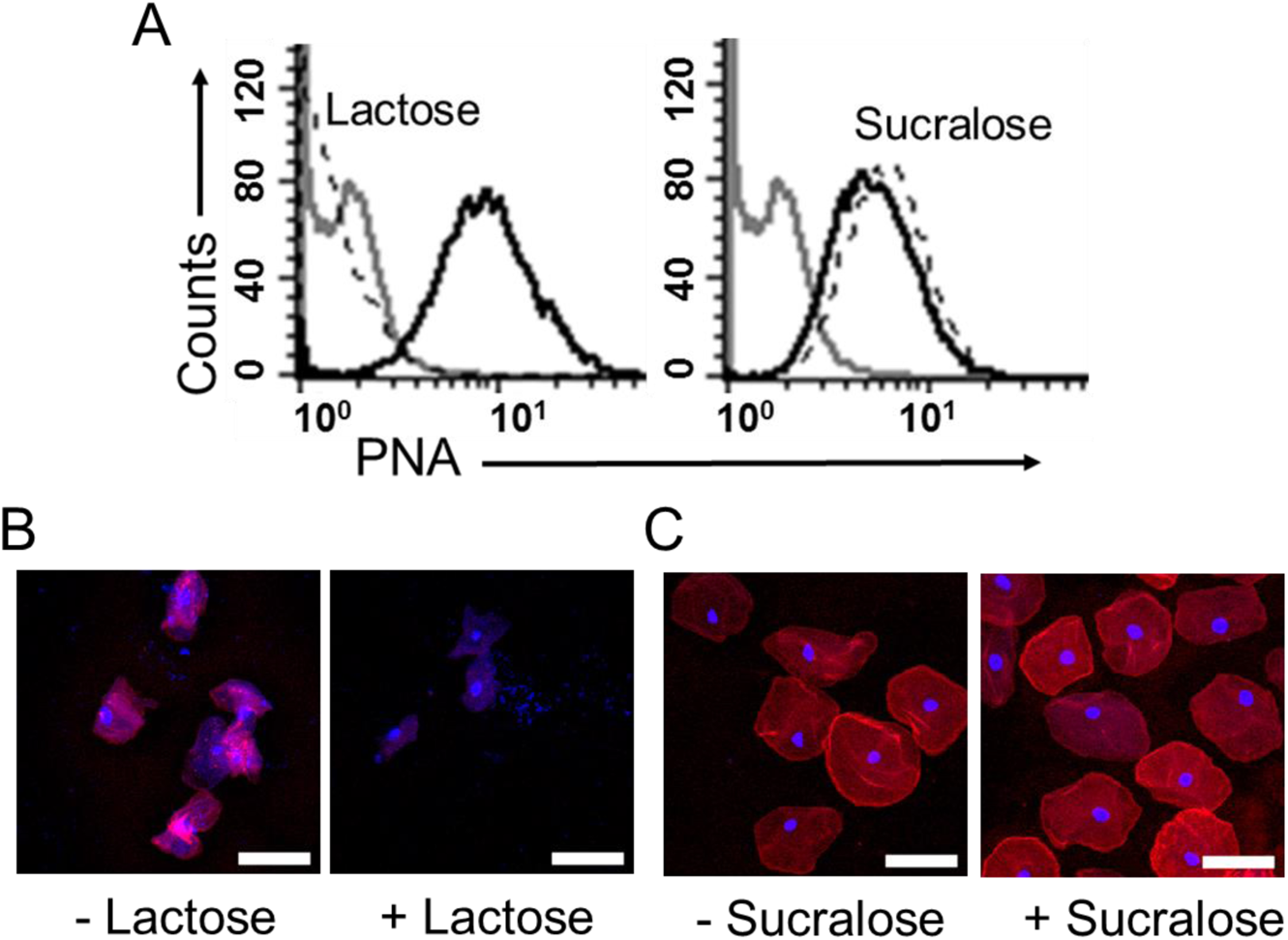
Verification of PNA selectivity for recognition of galactose containing glycans. VECs isolated from vaginal swabs of women with BV were treated with exogenous sialidase (*A.u.* sialidase), followed by staining with PNA-rhodamine in the presence of either 10mM lactose or sucralose. (**A**) Histograms show binding of PNA to VECs in the presence (dotted line) or absence (solid black line) of competing saccharide. Grey = Unstained VECs. (**B**) Representative confocal images of VECs stained with PNA in presence and absence of lactose. (**C**) Representative confocal images of VECs stained with PNA in presence and absence of sucralose. Red = PNA, Blue = DAPI. Scale bars = 50 µm.

**Fig. S6:**
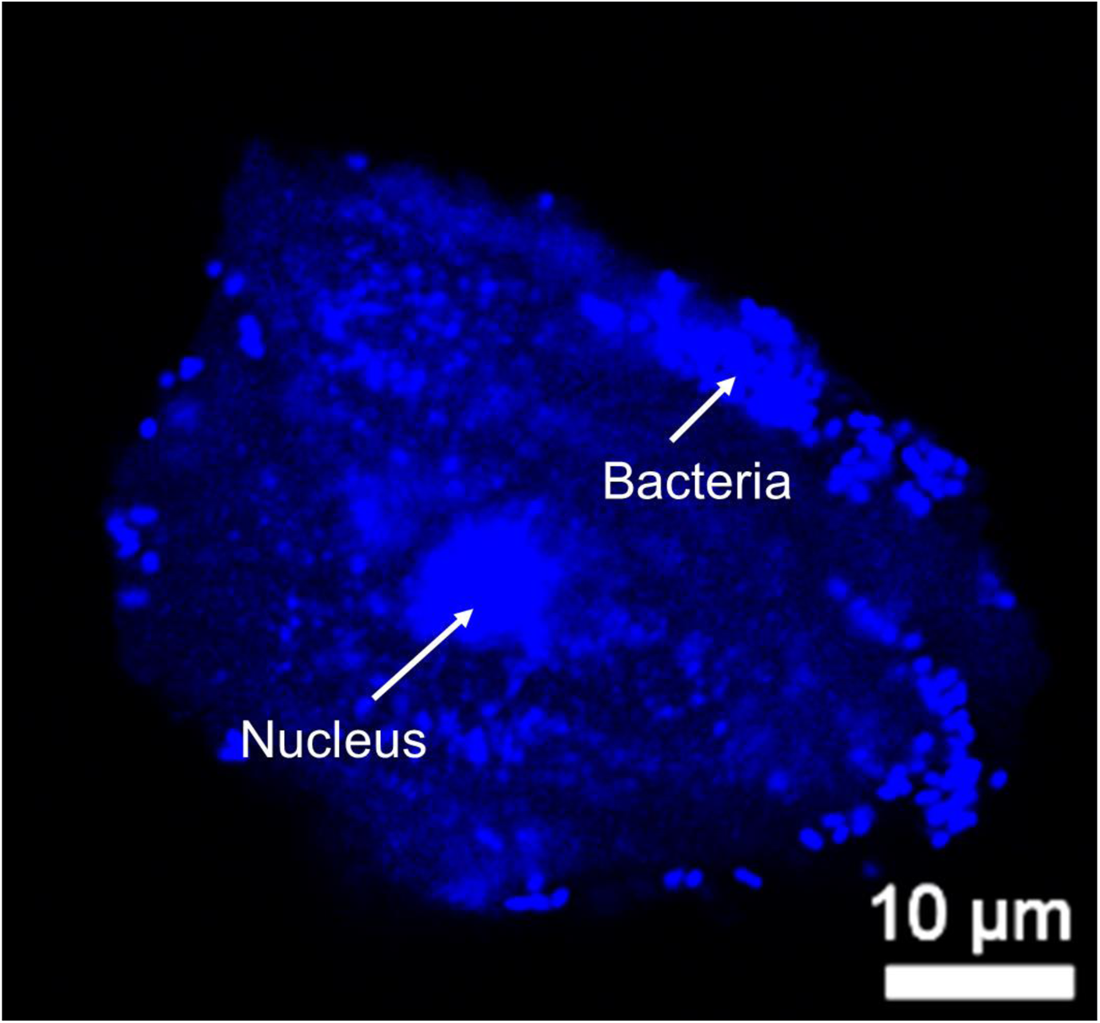
Visualization of bacteria attached to VECs (Clue cell phenotype). Confocal image of DAPI (Blue) stained VEC from a BV specimen. Nucleus is observed in the center. Blue puncta distributed around the nucleus and close to the cell membrane are bacterial cells stained with DAPI, giving a phenotype similar to that observed in wet mounts of women with BV.

**Fig. S7:**
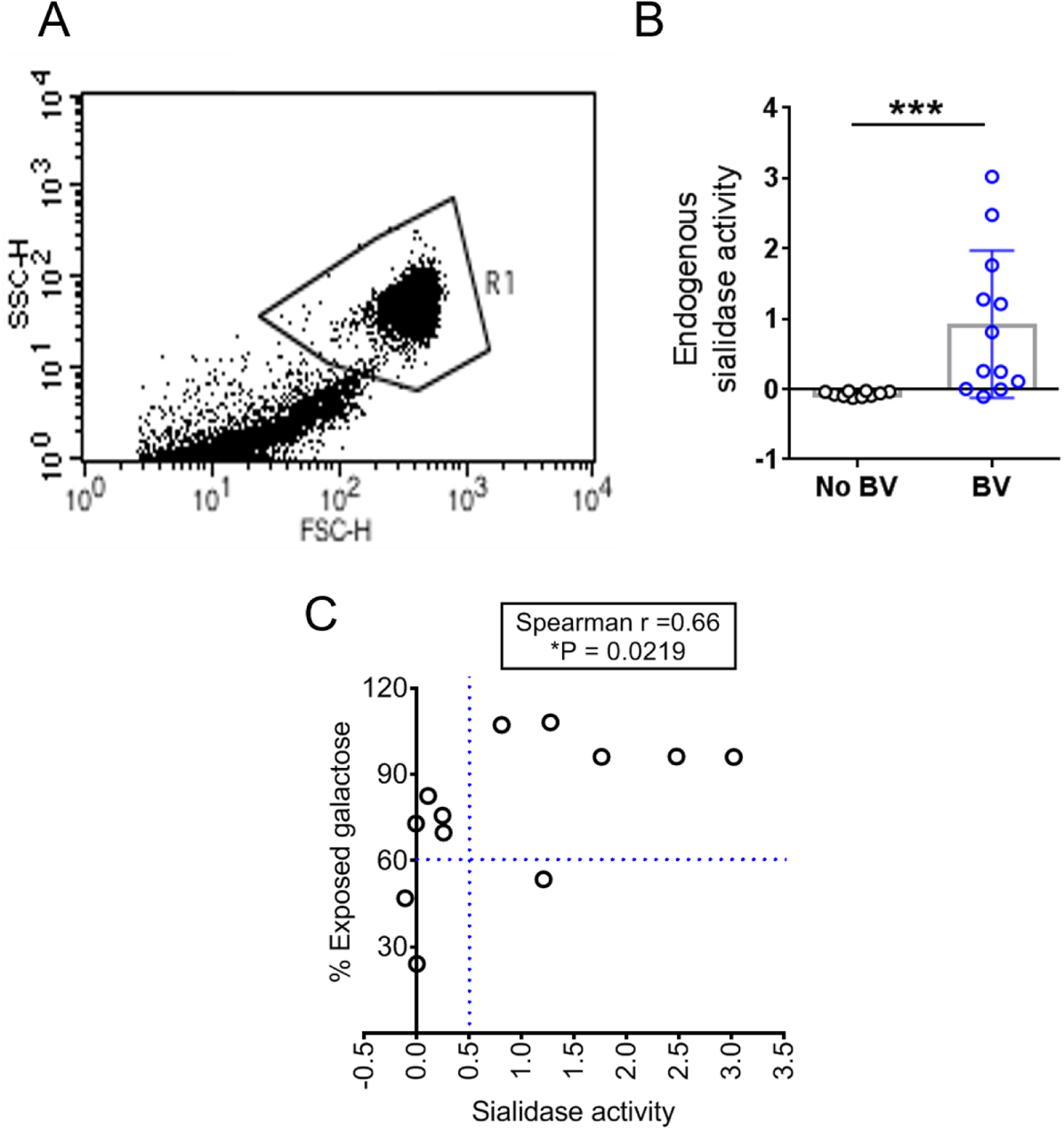
Galactose exposure on VECs correlates with sialidase activity in BV. (**A**) The dot plot shows flow cytometry gating strategy for selection of VECs based on forward (FSC-H) and side scatter (SSC-H). (**B**) Endogenous sialidase activity in individual vaginal swab eluates estimated using 4MU-Sia assay. (**C**) Graph shows correlation analysis for galactose exposure and sialidase activity observed in vaginal specimens of individual women. Samples that were pooled from multiple subjects were not included in this analysis.

**Fig. S8:**
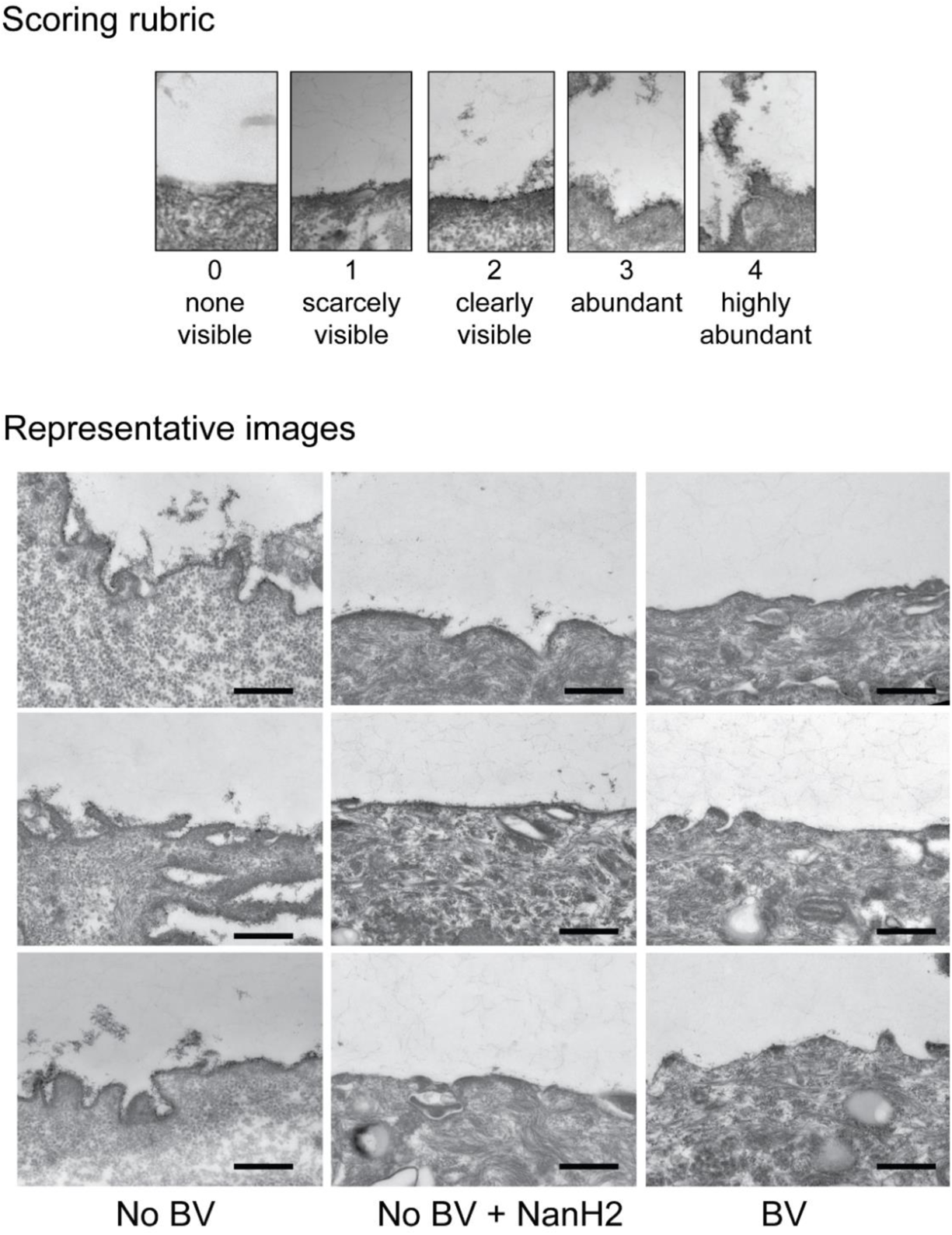
Rubric for vaginal epithelial glycocalyx scoring and representative images. Top: Rubric provided to three blinded observers. Bottom: Representative transmission electron microscopy (TEM) images of VECs acquired in a blinded fashion from (i) No BV specimens (left) - glycocalyx appears as a fuzzy layer close to the epithelial cell membrane, (ii) No BV specimens - treated with *Gardnerella* NanH2 sialidase (middle), and (iii) BV specimens (right). Images were acquired in a blinded fashion and scored by three observers blinded to the status of the sample. Scoring of the glycocalyx is shown in Figures 7D, E. All images were acquired at 25,000 X. Scale bars are 500 nm.

**Table S1.** Characteristics of the study population.

| **<u>Characteristic</u>** | **<u>Overall</u>** | **<u>BV</u>** | **<u>No BV</u>** | **<u>P-value</u>** |
| --- | --- | --- | --- | --- |
|  | N=258 | n=97 | n=161 |  |
| Age (mean years ± SD) | 25.7 ± 6.2 | 25.7 (0.6) | 25.7 (0.5) | 0.9519 |
| Race |  |  |  | <0.001 |
| Black | 130 (50.4%) | 66 (68.0%) | 64 (39.8%) |  |
| White | 111 (43.0%) | 27 (27.8%) | 84 (52.2%) |  |
| Other | 17 (6.6%) | 4 (4.1%) | 13 (8.1%) |  |
| Marital status |  |  |  | 0.7683 |
| Single, never married | 143 (55.4%) | 56 (57.7%) | 87 (54.0%) |  |
| Married or living with partner | 95 (36.8%) | 33 (34.0%) | 62 (38.5%) |  |
| Separated, divorced, or widowed | 20 (7.8%) | 8 (8.2%) | 12 (7.5%) |  |
| Education |  |  |  | <0.001 |
| ≤ High School | 85 (33%) | 43 (44.3%) | 42 (26.1%) | |
| Some college | 102 (39.5%) | 41 (42.3%) | 61 (37.9%) |  |
| College graduate | 71 (27.5%) | 13 (13.4%) | 58 (36.0%) |  |
| Nulligravid | 89 (34.5%) | 18 (18.6%) | 71 (44.1%) | <0.001 |
| Nulliparous | 122 (47.3%) | 32 (33.0%) | 90 (55.9%) | <0.001 |
| Sexually transmitted infection |  |  |  |  |
| *Chlamydia trachomatis* | 7 (2.8%) | 5 (5.2%) | 2 (1.3%) | 0.0644 |
| *Neisseria gonorrhoeae* | 3 (1.2%) | 3 (3.1%) | 0 | 0.0257 |
| *Trichomonas vaginalis* | 7 (2.7%) | 6 (6.3%) | 1 (0.6%) | 0.0080 |
| Prior diagnosis of sexually transmitted infection | 92 (36.7) | 42 (43.3%) | 50 (31.1%) | 0.0482 |
| Prior diagnosis of BV | 67 (26.0%) | 29 (29.9%) | 38 (23.6%) | 0.4920 |
| Currently pregnant | 32 (18.9%) | 16 (20.3%) | 16 (17.8%) | 0.6834 |
| Lifetime number of sexual partners (mean ± SD) | 8.6 ± 12.0 | 9.5 (1.4) | 8.1 (0.9) | 0.3424 |

## Methods

### Vaginal specimen source

Previously collected vaginal swabs leftover from the inaugural contraceptive CHOICE project at Washington University^102^ were used in this study. CHOICE received informed consent in writing from all participants at enrollment, as well as permission to use vaginal samples for future studies. This study was reviewed and approved by the Institutional Review Board at Washington University School of Medicine (IRB ID # 201108155). CHOICE inclusion criteria were: reported sexual activity in the past six months or anticipated sexual activity with a male partner and the desire to prevent pregnancy by use of contraception. Exclusion criteria were ages outside the 14-45 year age range and history of tubal ligation or hysterectomy, as previously described^102^.

### Specimen collection, handling, and Nugent scoring

Upon enrollment, women were given a double-headed rayon swab inside a collection tube (Starplex Scientific Inc, Etobicoke, Ontario, Canada) and were instructed on a self-collection protocol. Immediately after collection, a single technician rolled one swab onto a glass slide. Remaining swab material was stored at −80 °C. Slides were heat-fixed, Gram stained and subjected to Nugent scoring. In general, the Nugent scoring^66^ system uses weighted quantitation of bacterial morphotype to reflect the overall character of vaginal bacteria present. Nugent scores are reflected on a scale of 0 to 10 combined from three components for each woman: 1) a score of 0-4 reflecting the presence of rod-shaped Gram-positive lactobacilli where 0 indicates highest numbers, 2) a score of 0-4 reflecting the presence of Gram-negative and Gram-variable morphotypes where 4 indicates the highest numbers, and 3) a score of 0-2 reflecting the presence of curved rods. A woman with score of 0-3 is considered not to have BV (without BV/No BV, “normal”), and a woman with score of 7-10 is considered BV-positive.

### Specimen selection

A total of 258 specimens from individual women were used in the experiments reported here. Inclusion criteria for this sub-study were an available specimen Nugent score of 0-3 (No BV) or 7-10 (BV). Samples represent a cross-section of women from the same approximate (2 year) collection time window of CHOICE enrollment. Demographic characteristics of the participants in this sub study are presented in **Table S1**. We did not exclude specimens from women who were pregnant, menstruating, or those with sexually transmitted infections.

### Specimen processing

For all analyses of vaginal epithelial cells, vaginal swabs were selected based on Nugent score from reproductive age women with BV (Nugent score 7-10) or without BV (Nugent score 0-3). No attempts were made to exclude swabs from women who had clinical indications of sexually transmitted infection, although rates were very low. Vaginal swabs were stored at −80 °C until processed further. Swabs were cut to approximately ~3.5-4.0 cm and enclosed in a cryotube. Swabs were removed from the freezer onto wet ice and eluted by immersing in 1 mL of 100 mM sodium acetate (pH 5.5, swab elution buffer (SEB) incubated in an ice bucket for 1 h followed by shaking at 500 rpm, using a IKA MS 3 digital, for 10 min at room temperature. The individual swab eluates were transferred to microcentrifuge tubes and kept on ice. To check for sialidase activity, 50 µL of eluate was resuspended and transferred to 96 well round bottom black polypropylene plates. Sialidase assay was performed as described below. In parallel, a second elution was performed by again immersing the same swabs in 1 mL of SEB in individual cryo tubes, followed immediately by shaking at 500 rpm for 10 min at room temperature. Cells were isolated from the swab eluates by centrifugation at 1000 x g for 5 min at room temperature after each elution step. Microscopic examination of the isolated cells confirmed the morphology as polygonal squamous epithelial cells with a diameter of 50-70 µm and no other morphologies were observed.

Epithelial cells obtained from vaginal swabs were processed individually for detection of glycocalyx by transmission electron microscopy (TEM) and sialic acid analysis by MAL-II staining. For lectin analyses which required larger numbers of cells, like quantification of PNA binding by flow cytometry, VECs were pooled from multiple swabs in the same group (Nugent 0-3 or Nugent 7-10) to have sufficient packed cell pellet volume (~50 – 100 µL). For analytical studies focused on *N-* and *O*-glycan characterization, frozen VEC pellets were sent to the University of California San Diego GlycoAnalytics core. An additional selection criterion was applied to these samples: swabs in the BV group with detectable high sialidase activity (relative sialidase activity >3 for most specimens) as estimated by sialidase assay on the first swab eluate, were pooled together. For each study condition, approximately 100-200 µL of packed cell volume was obtained by pooling VECs isolated from 5 or 10 vaginal swab eluates in the same group.

### Fluorometric assay for sialidase activity

To measure sialidase activity, 50 µL of vaginal swab eluate was diluted 3-fold into 100 mM sodium acetate, pH 5.5, containing 200 µM 4-methylumbelliferyl-Neu5Ac (4-MU-Sialic acid) (Goldbio) in a 96 well round bottom black polypropylene plates and sealed with an optical clear film. Substrate hydrolysis was monitored using the fluorescence of 4-MU (Excitation 365 nm, Emission 440 nm) in a Tecan M200 plate reader every 2 min for 2 h at 37 °C.

### Preparation of *Gardnerella* sialidases NanH2 and NanH3

*Gardnerella* NanH2 and NanH3 (truncated versions lacking the transmembrane segments of both proteins and the signal sequence of NanH2), along with the vector control, were expressed in *E. coli* and purified by nickel chromatography as previously described^62^. In experiments with recombinant *Gardnerella* sialidases NanH2/NanH3, and sialidase from *Arthrobacter ureafaciens* (*A.u.* sialidase), enzyme activities were normalized as described earlier^62^. Similar amounts of enzyme activity were used to investigate their ability to cleave sialic acid from VECs.

### Visualization of glycocalyx on VECs by transmission electron microscopy (TEM)

VECs eluted from individual vaginal specimens of women with or without BV were washed with phosphate buffered saline (PBS) and processed for glycocalyx staining. For treatment of VECs with recombinant *Gardnerella* sialidase NanH2, cells obtained from individual specimens of women without BV were divided equally into two microcentrifuge tubes such that one half was treated with NanH2 in 300 µL SEB, and the other was mock treated with the same amount of buffer alone. Cells were kept at 37 °C on a rotor. After 1 h, cells were pelleted by centrifugation at 2000 g for 5 min and washed with 1 mL PBS twice. VECs were processed for glycocalyx detection using ruthenium red and osmium tetroxide (OsO_4_) as described by Fassel and Edmiston^65^. Briefly, VECs isolated from swab eluates were fixed and stained using 100 mM sodium cacodylate buffer (Polysciences Inc., Warrington, PA), pH 7.2, containing 0.15% ruthenium red, 2% paraformaldehyde and 2.5% glutaraldehyde for 1 h at RT. VECs were then washed with 100 mM cacodylate buffer containing 0.15% ruthenium red. Secondary fixation was performed with 1% OsO_4_ and 0.15% ruthenium red in 100 mM cacodylate buffer for 1 h (Polysciences Inc.). Samples were then rinsed extensively in deionized water (dH2O) prior to en bloc staining with 1% aqueous uranyl acetate for 1 h (Ted Pella Inc., Redding, CA). Following several rinses in dH20, samples were dehydrated in a graded series of ethanol and embedded in Eponate 12 resin (Ted Pella Inc.). Sections of 95 nm were cut with a Leica Ultracut UCT ultramicrotome (Leica Microsystems Inc., Bannockburn, IL), stained with uranyl acetate and lead citrate, and viewed on a JEOL 1200 EX transmission electron microscope (JEOL USA Inc., Peabody, MA) equipped with an AMT 8 megapixel digital camera and AMT Image Capture Engine V602 software (Advanced Microscopy Techniques, Woburn, MA). For glycocalyx scoring, the microscopist and scorers were blinded to the experimental treatment, as well as the BV status of the sample. 10 images of the epithelial cell membrane were acquired from each specimen at 25,000 X magnification. Images were labelled randomly to facilitate the blinded glycocalyx scoring. Images were scored between zero to five based on the abundance of glycocalyx, with zero being least abundant or nearly absent and five being highly abundant. Data from three independent scorers was combined to compute the average glycocalyx abundance score for each image.

### Lectin staining of vaginal epithelial cells

Cells were isolated from vaginal swab eluates of women with and without BV as described above. Cells were derived from individual women whenever possible; when cell numbers were insufficient for flow cytometry, cells from 2-5 individuals were pooled for analysis. Cells were washed three times with PBS containing 0.08% sodium azide (wash buffer). Prior to lectin staining, cell pellets for each sample (with ~50 µL packed cell volume) was re-suspended in 1 mL wash buffer, divided equally into 2 tubes and centrifuged at 2000 x g for 2 min. Supernatants were removed, pellets were resuspended in 300 µL of SEB (untreated controls) or SEB containing 15 mU of exogenous sialidase from *Arthrobacter ureafaciens* (*A.u.* sialidase) (EY Labs). Cells were incubated at 37 °C on a rotor to avoid clumping. After 1 h, cells were washed with 1 mL of lectin staining buffer (LSB) (10 mM HEPES, pH 7.5, 0.15 M sodium chloride and 0.1 mM calcium chloride). Cells were stained with peanut agglutinin (PNA) by incubating with 10 µg/mL of PNA-rhodamine (PNA-Rh) (Vector Lab) in 300 µL LSB in an ice bucket with intermittent mixing (every 10-15 min). After approximately 1 h, PNA stained cells were washed with LSB, re-suspended in 150 µL LSB and analyzed by flow cytometry. About 10,000 events were acquired for the same gated population for each sample (see **Fig. S7A**>). Untreated cells were used to assess auto fluorescence; PNA-Rh binding was analyzed in the FL-2 channel on a BD FACSCalibur flow cytometer. Exposed galactose was calculated as a percentage of PNA-binding mean fluorescence intensity (MFI) observed to untreated cells compared to the PNA-binding MFI to VECs from the same sample pretreated with *A.u.* sialidase.

For MAL-II staining, cells from individual specimens were incubated with 5 µg/mL of MAL-II biotin (Vector Laboratories) in 300 µL LSB in an ice bucket with intermittent mixing (every 10-15 min). After ~1 h, cells were washed with an avidin staining buffer (ASB) (1% BSA, 0.05% NaN_3_ in PBS). To detect biotin, cells were then stained with 2 µg/mL Neutravidin-OregonGreen 488 in 300 µL ASB for ~45 min at 4 °C degrees in an ice bucket. Cells stained with secondary only were included as control. Finally, the MAL-II stained cells and controls were washed with PBS and processed further for imaging.

For confocal microscopy, lectin-stained cells were re-suspended in PBS containing 600 nM DAPI (Invitrogen) and incubated in an ice bucket for 15 min. 10 µL of the cell suspension was then transferred to a microscope slide, covered with a cover slip and sealed with a transparent nail polish. Images were captured from at least three independent fields for each sample using LSM 880 confocal laser scanning microscope (Carl Zeiss) using 40 X objective lens with immersion oil.

### Detection of Lewis X (CD15) on VECs

Cells were isolated from vaginal swab eluates of women without BV as described above. Cells were washed three times with PBS containing 0.08% sodium azide (wash buffer). Prior to staining with anti-CD15, that recognizes Lewis X antigen, the cell pellet for each sample was resuspended in 1 mL wash buffer, divided equally into 2 tubes and centrifuged at 2000 x g for 2 min. Supernatants were removed, and the pellet was resuspended in 300 µL of SEB or SEB containing 15 mU of *A.u.* sialidase (EY Labs). Cells were incubated in this buffer at 37 °C on a rotor to avoid clumping. After 1 hour, cells were washed with 1 mL of LSB. To block non-specific binding, cells were incubated with 5 µg/mL of Fc Block (BD, 564220) in 600 µL of LSB for 15 min on a rotor at room temperature. Next, each specimen was divided equally into three tubes, 200 µL each, for staining with either 5.6 µg/mL secondary antibody alone (anti-mouse IgM-AlexaFluor488, Jackson ImmunoResearch) or 5 µg/mL of isotype control (mouse IgM, BD 555581) or 5 µg/mL of primary antibody (anti-Lewis X (Human CD15), BD 555400, clone HI98 (also known as HIM1)) followed by the secondary antibody. Cells stained with anti-Lewis X or isotype control were incubated in an ice bucket for 1 h with intermittent mixing (every 10-15 min). After washing the cells with 1 mL of LSB, a second staining step was performed in a similar manner by adding the secondary antibody to all the samples. After 40 min cells were washed two times with LSB, re-suspended in 150 µL LSB and analyzed by flow cytometry. Unstained cells were processed similarly in parallel without any antibody staining. About 10,000 events were acquired for each sample; anti-Lewis X binding was analyzed in the FL-1 channel on a BD FACSCalibur flow cytometer. Secondary alone showed little binding to the VECs and overlapped with unstained controls (data not shown). For confocal microscopy, samples were processed and imaged as described above.

### Preparation of crude cell lysate for glycan extraction

All analyses of VEC *N-* and *O-*glycans were performed at GlycoAnalytics Core at the University of California San Diego, California, USA. VECs isolated from vaginal swab (100 µL of packed cell pellet) were re-suspended in 300 µL of ultrapure water (Invitrogen) with 5 µL EDTA free protease inhibitor (EMD-Millipore Corp.) and sonicated in an ultrasonic water bath (Fisher Scientific Sonicator Model FS-9) until it formed a uniform suspension in water. Protein was quantified using BCA assay and further extraction of glycans was performed using a defined amount of protein as described in the methods below.

### Isolation and purification of *N*-glycans

*N*-linked glycans were isolated from glycoproteins in the crude cell lysate by overnight incubation with glycerol free recombinant PNGase-F (P0709S, NEB) at 37 °C. Briefly, cell extract for 500 µg of protein was mixed with a denaturing buffer (NEB kit) and boiled at 100 °C for 15 min. Proteins were solubilized by incubation with NP-40 for 1h at room temp, with intermittent vortexing every 15 min. *N*-glycans were extracted using PNGase-F and purified using a tandemly connected Sep-Pak C18 and porous graphitized carbon (PGC) cartridge equilibrated in water. PGC bound glycans were eluted with 30% acetonitrile containing 0.1% trifluoroacetic acid (TFA) in water and then lyophilized. The purified pool of glycans were processed further as required for each analysis.

### Isolation and purification of *O*-glycans

To release *O*-linked glycans by reductive β-elimination, crude cell lysate for 500 µg of protein was incubated with 0.05 M NaOH / 1 M NaBH_4_ at 45 °C for 16 h with constant stirring. Samples were then neutralized using ice-cold 30% aqueous acetic acid, followed by removal of sodium salts using cation exchange resin (Dowex 50Wx8, H^+^ Form, Bio-Rad 142-1451). The samples were co-evaporated using a 9:1 (v/v) methanol/acetic acid mixture and with absolute methanol to remove borate. The extracted *O*-glycans were purified using Sep-Pak C18 column, where the flow through from Sep-Pak C18 was lyophilized and used for further analysis^103^. Additionally, for linkage analysis by gas chromatography-mass spectrometry (GC-MS), charged *O*-linked glycans were separated (described below) from neutral *O*-glycans to reduce interference from contaminating neutral saccharides in the vaginal specimens.

### Sialic acid analysis on *N-* and *O*-glycans by reverse phase-liquid chromatography

For sialic acid analysis, glycans derived from 25 μg of protein extract were heated to 80 °C in 2 M acetic acid for 3 h. The released sialic acids were collected by ultrafiltration through a 3,000 MWCO filter and derivatized with 1,2-diamino-4,5-methylenedioxybenzene (DMB) (Sigma Aldrich), as described earlier^104^. The DMB-derivatized sialic acids were eluted isocratic with acetonitrile (8%) and methanol (7%) in Milli-Q^®^ water and analyzed either by HPLC using Acclaim 120 C18, 5 µM column (4.6 x 250 mm, Thermo-Dionex) at a flow rate of 0.9 mL/min over 30 min or using an Acquity™ UPLC system, with fluorescence detector set at λ_ex_ 373 nm, λ_em_ 448 nm. For UPLC, samples were eluted over 10 min using Acquity UPLC® BEH C18, 1.7 µM column (2.1 x 50 mm, Waters) at a flow rate of 0.2 mL/min. The DMB-derivatized sialic acids were identified and quantified by comparing elution times and peak areas to known standards using Chromeleon^®^ software. Data shown is from two independent experiments that were conducted ~three years apart. For the first experiment, VECs from 10 specimens were combined to obtain each pool as a greater biomass was required to complete all the analytical studies (shown in **Fig. 2–4**) on the same set of samples. The second experiment was conducted to specifically validate the preliminary findings of this experiment using a larger set of samples, for which VECs from 5 specimens were combined to obtain each pool.

### 2-Aminobenzamide (2-AB) labeling of purified N-glycans

The purified *N*-glycans were labeled with 2-AB as described earlier^105^. Briefly, samples were dissolved in 10 μL solution of 0.44 M 2-AB in acetic acid-DMSO mixture (35:65 v/v) containing 1 M sodium cyanoborohydride. Samples were then incubated at 65 °C for 2.5 h. 2-AB labeled glycans were purified using GlycoClean S cartridge (GLYKO) following the manufacturer’s protocol.

### 2-AB profiling with High-Performance Anion-Exchange Chromatography (HPAEC)-Fluorescence detection

Profiling of 2-AB labeled glycans was obtained using a Dionex CarboPac^®^ PA1 (4 X 250 mm) anion exchange column along with a guard column (4 X 50 mm) at flow rate of 1 mL/min. Glycans were separated in 100 mM sodium hydroxide with a sodium acetate gradient of 0-250 mM in 0-75 min. Data was collected using the Dionex ICS-3000 HPLC system with Ultimate 3000 fluorescence detector (Dionex) set at λ_ex_ 330 nm at λ_em_ 420 nm, with sensitivity 7. The data was processed using Chromeleon™ software (Thermo Scientific). Glycans from RNAse B and bovine fetuin were processed in parallel and labeled as known standards with high mannose and complex type *N*-glycans.

### Separation of *O*-glycans by charge

Anionic glycans were captured using liquid chromatography-Amino (LC-NH_2_) cartridges (Supelco, Part no. 504483, 1 mL). Briefly, total *O-*glycans (reconstituted in water after purification as described above) were loaded on the cartridge and the column was washed with 3 mL of ultrapure water (Invitrogen) which was collected as flow-through. Anionic species were eluted with 2 mL of 100 mM ammonium acetate and lyophilized.

### Per-*O*-methylation of glycans

Purified *O*-glycans, dried in glass reaction tubes, were dissolved in 350 µL of anhydrous DMSO by stirring for 1 h. To this solution was added a freshly prepared slurry (50 µL) of powdered NaOH in anhydrous DMSO, followed by immediate addition of 500 µL of methyl iodide. This reaction mixture was stirred vigorously at room temperature for 40 min. The reaction was quenched by adding 1 mL of ice-cold water followed by extraction of methylated glycans using chloroform. Finally, the chloroform was dried using a stream of nitrogen.

### MALDI-TOF profiling of *O*-glycans

Permethylated glycans were dissolved in absolute methanol and mixed 1:1 with super-DHB matrix solution and spotted on the matrix-assisted laser desorption/ionization (MALDI) plate. The samples were air-dried and analyzed in positive-reflectron mode on a MALDI-qTOF mass spectrometer (Applied Biosystems QSTAR)^106^. Structures for were manually annotated and verified using the glycobioinformatics tool GlycoWorkBench^107,108^. The proposed assignments for the selected peaks were based on the knowledge of mammalian glycan biosynthesis pathways. The proposed structures were confirmed using MS/MS and GC-MS linkage analysis as described below.

### MS/MS analysis of *O*-glycans

All spectra were acquired in the positive ionization mode over a range of 250-2000 amu on LTQ-XL Orbitrap Discovery Electrospray Ionization Mass Spectrometer (Thermo Scientific). Permethylated glycans were dissolved in absolute methanol, followed by addition of 1mM sodium hydroxide (NaOH), and direct injection into the mass spectrometer with a constant flow rate of 5 mL/min. The ion spray voltage was optimized at 3.7 kV with a sheath gas flow rate 12 mL/min. All MS and MS(n) spectra were collected in positive ionization mode with resolution of 30000. The other instrument parameters for positive-ion detection were: capillary temperature 145 °C; capillary voltage 47 V. Tandem mass spectrometry (MSn) was performed via collision-induced dissociation (CID) on selected parent ions with collision energy of 15-20%.

### Glycan linkage analysis by GC-MS

To determine linkage positions of vaginal epithelial *O*-glycans, permethylated glycans (permethylation described above) were hydrolyzed with 4 N TFA at 100 °C for 4 h, followed by removal of acid using nitrogen flush. The sample was then reduced using ~5 mg sodium borodeuteride (NaBD_4_) solution in 1 M ammonium hydroxide, and acetylated using pyridine and acetic anhydride (1:1 v/v) mixture at 100 °C for 1 h^109^. Partially methylated acetylated alditol (PMAA) derivatives were analyzed by GC-MS^72,110^ using Restek-5ms capillary column (30 m x 0.25 mm x 0.25 µm) on an Agilent Technologies 7820A GC System 5975 Series Gas Mass Selective Detector (GC-MS). Identifications are achieved by using a combination of retention times and the mass fragmentation pattern in EI-MS mode.

### Lectin analysis of VECs treated with *Gardnerella* sialidases

Prior to lectin staining, the cell pellet for each pooled sample (with ~100 µL of packed cell volume) was re-suspended in 1 mL wash buffer, divided equally into 4 tubes and centrifuged at 2000 x g for 2 min. Supernatants were removed, and the pellet was resuspended in 300 µL of SEB or SEB containing either similar amounts of enzyme activity of NanH2, NanH3, *A.u.* sialidase or similar volume of preparation from vector control. Cells were incubated in this buffer at 37 °C on a rotor to avoid clumping. After 1 h, supernatants were collected for sialic acid analysis by DMB-HPLC, and cells were washed with 1 mL of LSB. For staining with PNA, cells were incubated with 5 µg/mL of PNA-AlexaFluor647 (AF647) in 300 µL LSB in an ice bucket with intermittent mixing (every 10-15 min). After approximately 1h, PNA stained cells were washed with LSB, re-suspended in 150 µL LSB and analyzed by flow cytometry. About 10,000 events were acquired for each sample; PNA-AF647 binding was analyzed in the FL-4 channel on a BD FACSCalibur flow cytometer. For confocal microscopy, lectin-stained cells were processed for imaging as described above.

## Data Availability

All data produced in the present work are contained in the manuscript

## Acknowledgements

We are grateful to Dr. Wandy L. Beatty and the staff of the Molecular Microbiology Imaging Facility at Washington University in St. Louis (MO, USA) for assistance with transmission electron microscopy, to Dr. Sulabha Argade and Dr. Mousumi Paulchakrabarti at the GlycoAnalytics Core, Glycobiology Research and Training Center, University of California San Diego (CA, USA) for expert technical assistance with glycan characterization. We also acknowledge Dr. Jeffrey F. Peipert for leadership of the inaugural Contraceptive CHOICE project from which specimens were originally archived, and Jennifer Reed (formerly Bick) for technical assistance with Nugent scoring of clinical specimens. Finally, we thank all of the participants who generously shared samples used in this study. This work was funded by the National Institute of Allergy and Infectious Diseases (R01 AI114635 to ALL and WGL, R01 AI127554 to WGL), the Burroughs Wellcome Fund Preterm Birth Initiative (to ALL). This work was also supported by the Stephen I. Morse fellowship to K.A. (Department of Molecular Microbiology, Washington University in St. Louis, MO, USA). Funding for the Contraceptive CHOICE Project was provided by an anonymous foundation.

## Author contributions

A.L.L., and W.G.L. conceived and supervised the project. K.A., B.C., A.L.L., and W.G.L. designed the experiments; K.A. performed enzyme assays, imaging, flow cytometry, and HPLC data acquisition; B.C. acquired mass spectrometric and HPAEC data; L.S.R. expressed and purified NanH2 and NanH3 proteins; K.A., L.S.R., and W.G.L performed blinded scoring of electron micrographs; J.E.A. provided Nugent scoring data and performed analysis of demographic/clinical characteristics; K.A., B.C., A.L.L., and W.G.L. performed data analysis; K.A., A.L.L., and W.G.L. made figures, and wrote the manuscript with input/approval from all the others.

## References

1. Callahan, B. J. et al. Replication and refinement of a vaginal microbial signature of preterm birth in two racially distinct cohorts of US women. Proc Natl Acad Sci U S A 114, 9966–9971, doi:10.1073/pnas.1705899114 (2017).

2. Hillier, S. L., Bernstein, K. T. & Aral, S. A Review of the Challenges and Complexities in the Diagnosis, Etiology, Epidemiology, and Pathogenesis of Pelvic Inflammatory Disease. J Infect Dis 224, S23–S28, doi:10.1093/infdis/jiab116 (2021).

3. Witkin, S. S. & Linhares, I. M. Why do lactobacilli dominate the human vaginal microbiota? BJOG 124, 606–611, doi:10.1111/1471-0528.14390 (2017).

4. Kroon, S. J., Ravel, J. & Huston, W. M. Cervicovaginal microbiota, women’s health, and reproductive outcomes. Fertil Steril 110, 327–336, doi:10.1016/j.fertnstert.2018.06.036 (2018).

5. Amabebe, E. & Anumba, D. O. C. The Vaginal Microenvironment: The Physiologic Role of Lactobacilli. Front Med (Lausanne) 5, 181, doi:10.3389/fmed.2018.00181 (2018).

6. Al-Memar, M. et al. The association between vaginal bacterial composition and miscarriage: a nested case-control study. BJOG 127, 264–274, doi:10.1111/1471-0528.15972 (2020).

7. Piot, P., Van Dyck, E., Godts, P. & Vanderheyden, J. The vaginal microbial flora in non-specific vaginitis. Eur J Clin Microbiol 1, 301–306, doi:10.1007/BF02019976 (1982).

8. Hill, G. B., Eschenbach, D. A. & Holmes, K. K. Bacteriology of the vagina. Scand J Urol Nephrol Suppl 86, 23–39 (1984).

9. Hillier, S. L., Krohn, M. A., Rabe, L. K., Klebanoff, S. J. & Eschenbach, D. A. The normal vaginal flora, H2O2-producing lactobacilli, and bacterial vaginosis in pregnant women. Clin Infect Dis 16 Suppl 4, S273–281, doi:10.1093/clinids/16.supplement_4.s273 (1993).

10. Fredricks, D. N., Fiedler, T. L. & Marrazzo, J. M. Molecular identification of bacteria associated with bacterial vaginosis. N Engl J Med 353, 1899–1911, doi:10.1056/NEJMoa043802 (2005).

11. Ravel, J. et al. Vaginal microbiome of reproductive-age women. Proc Natl Acad Sci U S A 108 Suppl 1, 4680–4687, doi:10.1073/pnas.1002611107 (2011).

12. Hay, P. E. et al. Abnormal bacterial colonisation of the genital tract and subsequent preterm delivery and late miscarriage. BMJ 308, 295–298, doi:10.1136/bmj.308.6924.295 (1994).

13. Leitich, H. et al. Antibiotic treatment of bacterial vaginosis in pregnancy: a meta-analysis. Am J Obstet Gynecol 188, 752–758, doi:10.1067/mob.2003.167 (2003).

14. Holst, E., Goffeng, A. R. & Andersch, B. Bacterial vaginosis and vaginal microorganisms in idiopathic premature labor and association with pregnancy outcome. J Clin Microbiol 32, 176–186, doi:10.1128/jcm.32.1.176-186.1994 (1994).

15. Hillier, S. L. et al. Association between bacterial vaginosis and preterm delivery of a low-birth-weight infant. The Vaginal Infections and Prematurity Study Group. N Engl J Med 333, 1737–1742, doi:10.1056/NEJM199512283332604 (1995).

16. Fettweis, J. M. et al. The vaginal microbiome and preterm birth. Nat Med 25, 1012–1021, doi:10.1038/s41591-019-0450-2 (2019).

17. Soper, D. E. Bacterial vaginosis and surgical site infections. Am J Obstet Gynecol 222, 219–223, doi:10.1016/j.ajog.2019.09.002 (2020).

18. Wiesenfeld, H. C. et al. Lower genital tract infection and endometritis: insight into subclinical pelvic inflammatory disease. Obstet Gynecol 100, 456–463, doi:10.1016/s0029-7844(02)02118-x (2002).

19. Taylor, B. D., Darville, T. & Haggerty, C. L. Does bacterial vaginosis cause pelvic inflammatory disease? Sex Transm Dis 40, 117–122, doi:10.1097/OLQ.0b013e31827c5a5b (2013).

20. Ravel, J., Moreno, I. & Simon, C. Bacterial vaginosis and its association with infertility, endometritis, and pelvic inflammatory disease. Am J Obstet Gynecol 224, 251–257, doi:10.1016/j.ajog.2020.10.019 (2021).

21. Brotman, R. M. et al. Bacterial vaginosis assessed by gram stain and diminished colonization resistance to incident gonococcal, chlamydial, and trichomonal genital infection. J Infect Dis 202, 1907–1915, doi:10.1086/657320 (2010).

22. Bautista, C. T. et al. Association of Bacterial Vaginosis With *Chlamydia* and *Gonorrhea* Among Women in the U.S. Army. Am J Prev Med 52, 632–639, doi:10.1016/j.amepre.2016.09.016 (2017).

23. Atashili, J., Poole, C., Ndumbe, P. M., Adimora, A. A. & Smith, J. S. Bacterial vaginosis and HIV acquisition: a meta-analysis of published studies. AIDS 22, 1493–1501, doi:10.1097/QAD.0b013e3283021a37 (2008).

24. Vodstrcil, L. A., Muzny, C. A., Plummer, E. L., Sobel, J. D. & Bradshaw, C. S. Bacterial vaginosis: drivers of recurrence and challenges and opportunities in partner treatment. BMC Med 19, 194, doi:10.1186/s12916-021-02077-3 (2021).

25. Cook, R. L., Reid, G., Pond, D. G., Schmitt, C. A. & Sobel, J. D. Clue cells in bacterial vaginosis: immunofluorescent identification of the adherent gram-negative bacteria as *Gardnerella vaginalis*. J Infect Dis 160, 490–496, doi:10.1093/infdis/160.3.490 (1989).

26. Gardner, H. L. & Dukes, C. D. *Haemophilus vaginalis* vaginitis: a newly defined specific infection previously classified non-specific vaginitis. Am J Obstet Gynecol 69, 962–976 (1955).

27. Swidsinski, A. et al. Adherent biofilms in bacterial vaginosis. Obstet Gynecol 106, 1013–1023, doi:10.1097/01.AOG.0000183594.45524.d2 (2005).

28. Amsel, R. et al. Nonspecific vaginitis. Diagnostic criteria and microbial and epidemiologic associations. Am J Med 74, 14–22, doi:10.1016/0002-9343(83)91112-9 (1983).

29. Schwebke, J. R., Marrazzo, J., Beelen, A. P. & Sobel, J. D. A Phase 3, Multicenter, Randomized, Double-Blind, Vehicle-Controlled Study Evaluating the Safety and Efficacy of Metronidazole Vaginal Gel 1.3% in the Treatment of Bacterial Vaginosis. Sex Transm Dis 42, 376–381, doi:10.1097/OLQ.0000000000000300 (2015).

30. Srinivasan, S. et al. More than meets the eye: associations of vaginal bacteria with gram stain morphotypes using molecular phylogenetic analysis. PLoS One 8, e78633, doi:10.1371/journal.pone.0078633 (2013).

31. Hardy, L. et al. Unravelling the Bacterial Vaginosis-Associated Biofilm: A Multiplex *Gardnerella vaginalis* and *Atopobium vaginae* Fluorescence In Situ Hybridization Assay Using Peptide Nucleic Acid Probes. PLoS One 10, e0136658, doi:10.1371/journal.pone.0136658 (2015).

32. Amegashie, C. P. et al. Relationship between Nugent score and vaginal epithelial exfoliation. PLoS One 12, e0177797, doi:10.1371/journal.pone.0177797 (2017).

33. Roselletti, E. et al. Apoptosis of vaginal epithelial cells in clinical samples from women with diagnosed bacterial vaginosis. Sci Rep 10, 1978, doi:10.1038/s41598-020-58862-2 (2020).

34. Ceralli, F., Familiari, G., Marinozzi, G. & Muccioli-Casadei, D. The glycocalyx of the epithelial cells of the colon, observed in normal and ulcerous colitic conditions. Experientia 32, 1542–1544, doi:10.1007/BF01924441 (1976).

35. Levin, S. & Richter, W. R. Ultrastructure of cell surface coat (glycocalyx) in rat urinary bladder epithelium. Cell Tissue Res 158, 281–283, doi:10.1007/BF00219966 (1975).

36. Martins M. d. F. & Bairos, V. A. Glycocalyx of lung epithelial cells. Int Rev Cytol 216, 131–173, doi:10.1016/s0074-7696(02)16005-0 (2002).

37. Poole, J., Day, C. J., von Itzstein, M., Paton, J. C. & Jennings, M. P. Glycointeractions in bacterial pathogenesis. Nat Rev Microbiol 16, 440–452, doi:10.1038/s41579-018-0007-2 (2018).

38. Cohen, M. et al. Influenza A penetrates host mucus by cleaving sialic acids with neuraminidase. Virol J 10, 321, doi:10.1186/1743-422X-10-321 (2013).

39. Argueso, P., Woodward, A. M. & AbuSamra, D. B. The Epithelial Cell Glycocalyx in Ocular Surface Infection. Front Immunol 12, 729260, doi:10.3389/fimmu.2021.729260 (2021).

40. Arabyan, N. et al. Salmonella Degrades the Host Glycocalyx Leading to Altered Infection and Glycan Remodeling. Sci Rep 6, 29525, doi:10.1038/srep29525 (2016).

41. Cobb, B. A. & Kasper, D. L. Coming of age: carbohydrates and immunity. Eur J Immunol 35, 352–356, doi:10.1002/eji.200425889 (2005).

42. Varki, A. & Gagneux, P. Multifarious roles of sialic acids in immunity. Ann N Y Acad Sci 1253, 16–36, doi:10.1111/j.1749-6632.2012.06517.x (2012).

43. Dias, A. M. et al. Glycans as critical regulators of gut immunity in homeostasis and disease. Cell Immunol 333, 9–18, doi:10.1016/j.cellimm.2018.07.007 (2018).

44. Brazil, J. C. & Parkos, C. A. Finding the sweet spot: glycosylation mediated regulation of intestinal inflammation. Mucosal Immunol, doi:10.1038/s41385-021-00466-8 (2021).

45. Pinho, S. S. & Reis, C. A. Glycosylation in cancer: mechanisms and clinical implications. Nat Rev Cancer 15, 540–555, doi:10.1038/nrc3982 (2015).

46. Reily, C., Stewart, T. J., Renfrow, M. B. & Novak, J. Glycosylation in health and disease. Nat Rev Nephrol 15, 346–366, doi:10.1038/s41581-019-0129-4 (2019).

47. Lewis, A. L. & Lewis, W. G. Host sialoglycans and bacterial sialidases: a mucosal perspective. Cell Microbiol 14, 1174–1182, doi:10.1111/j.1462-5822.2012.01807.x (2012).

48. Yurewicz, E. C., Matsuura, F. & Moghissi, K. S. Structural characterization of neutral oligosaccharides of human midcycle cervical mucin. J Biol Chem 257, 2314–2322 (1982).

49. Yurewicz, E. C., Matsuura, F. & Moghissi, K. S. Structural studies of sialylated oligosaccharides of human midcycle cervical mucin. J Biol Chem 262, 4733–4739 (1987).

50. Andersch-Bjorkman, Y., Thomsson, K. A., Holmen Larsson, J. M., Ekerhovd, E. & Hansson, G. C. Large scale identification of proteins, mucins, and their *O*-glycosylation in the endocervical mucus during the menstrual cycle. Mol Cell Proteomics 6, 708–716, doi:10.1074/mcp.M600439-MCP200 (2007).

51. Moncla, B. J., Chappell, C. A., Debo, B. M. & Meyn, L. A. The Effects of Hormones and Vaginal Microflora on the Glycome of the Female Genital Tract: Cervical-Vaginal Fluid. PLoS One 11, e0158687, doi:10.1371/journal.pone.0158687 (2016).

52. Moncla, B. J. et al. Impact of bacterial vaginosis, as assessed by Nugent criteria and hormonal status on glycosidases and lectin binding in cervicovaginal lavage samples. PLoS One 10, e0127091, doi:10.1371/journal.pone.0127091 (2015).

53. Wang, L. et al. Studying the effects of reproductive hormones and bacterial vaginosis on the glycome of lavage samples from the cervicovaginal cavity. PLoS One 10, e0127021, doi:10.1371/journal.pone.0127021 (2015).

54. Briselden, A. M., Moncla, B. J., Stevens, C. E. & Hillier, S. L. Sialidases (neuraminidases) in bacterial vaginosis and bacterial vaginosis-associated microflora. J Clin Microbiol 30, 663–666, doi:10.1128/jcm.30.3.663-666.1992 (1992).

55. Howe, L. et al. Mucinase and sialidase activity of the vaginal microflora: implications for the pathogenesis of preterm labour. Int J STD AIDS 10, 442–447, doi:10.1258/0956462991914438 (1999).

56. Lewis, W. G. et al. Hydrolysis of secreted sialoglycoprotein immunoglobulin A (IgA) in *ex vivo* and biochemical models of bacterial vaginosis. J Biol Chem 287, 2079–2089, doi:10.1074/jbc.M111.278135 (2012).

57. Olmsted, S. S., Meyn, L. A., Rohan, L. C. & Hillier, S. L. Glycosidase and proteinase activity of anaerobic gram-negative bacteria isolated from women with bacterial vaginosis. Sex Transm Dis 30, 257–261, doi:10.1097/00007435-200303000-00016 (2003).

58. Kampan, N. C. et al. Evaluation of BV((R)) Blue Test Kit for the diagnosis of bacterial vaginosis. Sex Reprod Healthc 2, 1–5, doi:10.1016/j.srhc.2010.11.002 (2011).

59. Zhang, X. et al. [Relationship between vaginal sialidase bacteria vaginosis and chorioammionitis]. Zhonghua Fu Chan Ke Za Zhi 37, 588–590 (2002).

60. Cauci, S. & Culhane, J. F. High sialidase levels increase preterm birth risk among women who are bacterial vaginosis-positive in early gestation. Am J Obstet Gynecol 204, 142 e141–149, doi:10.1016/j.ajog.2010.08.061 (2011).

61. Lewis, W. G., Robinson, L. S., Gilbert, N. M., Perry, J. C. & Lewis, A. L. Degradation, foraging, and depletion of mucus sialoglycans by the vagina-adapted Actinobacterium *Gardnerella vaginalis*. J Biol Chem 288, 12067–12079, doi:10.1074/jbc.M113.453654 (2013).

62. Robinson, L. S., Schwebke, J., Lewis, W. G. & Lewis, A. L. Identification and characterization of NanH2 and NanH3, enzymes responsible for sialidase activity in the vaginal bacterium *Gardnerella vaginalis*. J Biol Chem 294, 5230–5245, doi:10.1074/jbc.RA118.006221 (2019).

63. Srinivasan, S. et al. Metabolic signatures of bacterial vaginosis. mBio 6, doi:10.1128/mBio.00204-15 (2015).

64. Luft, J. H. Fine structures of capillary and endocapillary layer as revealed by ruthenium red. Fed Proc 25, 1773–1783 (1966).

65. Fassel, T. A. & Edmiston, C. E., Jr. Ruthenium red and the bacterial glycocalyx. Biotech Histochem 74, 194–212, doi:10.3109/10520299909047974 (1999).

66. Nugent, R. P., Krohn, M. A. & Hillier, S. L. Reliability of diagnosing bacterial vaginosis is improved by a standardized method of gram stain interpretation. J Clin Microbiol 29, 297–301, doi:10.1128/jcm.29.2.297-301.1991 (1991).

67. Geisler, C. & Jarvis, D. L. Effective glycoanalysis with *Maackia amurensis* lectins requires a clear understanding of their binding specificities. Glycobiology 21, 988–993, doi:10.1093/glycob/cwr080 (2011).

68. Gagneux, P., Aebi, M., Varki, A. Evolution of Glycan Diversity. In Essentials of Glycobiology (Third edition). Editors: Varki, A., Cummings, RD., Esko, JD., et al. Cold Spring Harbor (NY): Cold Spring Harbor Laboratory Press; doi: 10.1101/glycobiology.3e.020 (2017).

69. Van den Steen, P., Rudd, P. M., Dwek, R. A. & Opdenakker, G. Concepts and principles of *O*-linked glycosylation. Crit Rev Biochem Mol Biol 33, 151–208, doi:10.1080/10409239891204198 (1998).

70. Brockhausen, I. & Stanley, P. *O*-GalNAc Glycans. In Essentials of Glycobiology (Third edition). Editors: Varki, A., Cummings, RD., Esko, JD. et al. Cold Spring Harbor (NY): Cold Spring Harbor Laboratory Press; doi: 10.1101/glycobiology.3e.010 (2017).

71. Domon, B. & Costello, C. E. A Systematic Nomenclature for Carbohydrate Fragmentations in Fab-Ms Ms Spectra of Glycoconjugates. Glycoconjugate J 5, 397–409, doi:Doi 10.1007/Bf01049915 (1988).

72. Bjorndal, H., Hellerqv. Cg, Lindberg, B. & Svensson, S. Gas-Liquid Chromatography and Mass Spectrometry in Methylation Analysis of Polysaccharides. Angew Chem Int Edit 9, 610-&, doi:DOI 10.1002/anie.197006101 (1970).

73. Hoskins, L. C. Human enteric population ecology and degradation of gut mucins. Dig Dis Sci 26, 769–772, doi:10.1007/BF01309606 (1981).

74. Yudin, A. I., Treece, C. A., Tollner, T. L., Overstreet, J. W. & Cherr, G. N. The carbohydrate structure of DEFB126, the major component of the cynomolgus Macaque sperm plasma membrane glycocalyx. J Membr Biol 207, 119–129, doi:10.1007/s00232-005-0806-z (2005).

75. Chacko, B. K. & Appukuttan, P. S. Peanut (*Arachis hypogaea*) lectin recognizes alpha-linked galactose, but not N-acetyl lactosamine in N-linked oligosaccharide terminals. Int J Biol Macromol 28, 365–371, doi:10.1016/s0141-8130(01)00139-8 (2001).

76. Pacheco, A. R. et al. Fucose sensing regulates bacterial intestinal colonization. Nature 492, 113–117, doi:10.1038/nature11623 (2012).

77. Goto, Y. et al. Innate lymphoid cells regulate intestinal epithelial cell glycosylation. Science 345, 1254009, doi:10.1126/science.1254009 (2014).

78. Nita-Lazar, M. et al. Desialylation of airway epithelial cells during influenza virus infection enhances pneumococcal adhesion via galectin binding. Mol Immunol 65, 1–16, doi:10.1016/j.molimm.2014.12.010 (2015).

79. Jeffries, J. L. et al. *Pseudomonas aeruginosa* pyocyanin modulates mucin glycosylation with sialyl-Lewis(x) to increase binding to airway epithelial cells. Mucosal Immunol 9, 1039–1050, doi:10.1038/mi.2015.119 (2016).

80. Kudelka, M. R., Stowell, S. R., Cummings, R. D. & Neish, A. S. Intestinal epithelial glycosylation in homeostasis and gut microbiota interactions in IBD. Nat Rev Gastroenterol Hepatol 17, 597–617, doi:10.1038/s41575-020-0331-7 (2020).

81. Paul, K., Boutain, D., Manhart, L. & Hitti, J. Racial disparity in bacterial vaginosis: the role of socioeconomic status, psychosocial stress, and neighborhood characteristics, and possible implications for preterm birth. Soc Sci Med 67, 824–833, doi:10.1016/j.socscimed.2008.05.017 (2008).

82. Borgogna, J. C. et al. Vaginal microbiota of American Indian women and associations with measures of psychosocial stress. PLoS One 16, e0260813, doi:10.1371/journal.pone.0260813 (2021).

83. Eylar, E. H., Madoff, M. A., Brody, O. V. & Oncley, J. L. The contribution of sialic acid to the surface charge of the erythrocyte. J Biol Chem 237, 1992–2000 (1962).

84. Gelberg, H., Healy, L., Whiteley, H., Miller, L. A. & Vimr, E. *In vivo* enzymatic removal of alpha 2-->6-linked sialic acid from the glomerular filtration barrier results in podocyte charge alteration and glomerular injury. Lab Invest 74, 907–920 (1996).

85. Hebbel, R. P. et al. Abnormal adherence of sickle erythrocytes to cultured vascular endothelium: possible mechanism for microvascular occlusion in sickle cell disease. J Clin Invest 65, 154–160, doi:10.1172/JCI109646 (1980).

86. Niculovic, K. M. et al. Podocyte-Specific Sialylation-Deficient Mice Serve as a Model for Human FSGS. J Am Soc Nephrol 30, 1021–1035, doi:10.1681/ASN.2018090951 (2019).

87. Sen, P., Ghosh, D. & Sarkar, C. Erythrocytic membrane anionic charge, sialic acid content, and their correlations with urinary glycosaminoglycans in preeclampsia and eclampsia. Scand J Clin Lab Invest 80, 343–347, doi:10.1080/00365513.2020.1750687 (2020).

88. Vimr, E. R. & Troy, F. A. Identification of an inducible catabolic system for sialic acids (nan) in *Escherichia coli*. J Bacteriol 164, 845–853, doi:10.1128/jb.164.2.845-853.1985 (1985).

89. Agarwal, K. & Lewis, A. L. Vaginal sialoglycan foraging by *Gardnerella vaginalis*: mucus barriers as a meal for unwelcome guests? Glycobiology 31, 667–680, doi:10.1093/glycob/cwab024 (2021).

90. Agarwal, K. et al. Glycan cross-feeding supports mutualism between *Fusobacterium* and the vaginal microbiota. PLoS Biol 18, e3000788, doi:10.1371/journal.pbio.3000788 (2020).

91. Coppenhagen-Glazer, S. et al. Fap2 of *Fusobacterium nucleatum* is a galactose-inhibitable adhesin involved in coaggregation, cell adhesion, and preterm birth. Infect Immun 83, 1104–1113, doi:10.1128/IAI.02838-14 (2015).

92. Wu, C. et al. Genetic and molecular determinants of polymicrobial interactions in *Fusobacterium nucleatum*. Proc Natl Acad Sci U S A 118, doi:10.1073/pnas.2006482118 (2021).

93. Okumura, C. Y., Baum, L. G. & Johnson, P. J. Galectin-1 on cervical epithelial cells is a receptor for the sexually transmitted human parasite *Trichomonas vaginalis*. Cell Microbiol 10, 2078–2090, doi:10.1111/j.1462-5822.2008.01190.x (2008).

94. Fichorova, R. N. et al. *Trichomonas vaginalis* Lipophosphoglycan Exploits Binding to Galectin-1 and -3 to Modulate Epithelial Immunity. J Biol Chem 291, 998–1013, doi:10.1074/jbc.M115.651497 (2016).

95. Tecle, E., Reynoso, H. S., Wang, R. & Gagneux, P. The female reproductive tract contains multiple innate sialic acid-binding immunoglobulin-like lectins (Siglecs) that facilitate sperm survival. J Biol Chem 294, 11910–11919, doi:10.1074/jbc.RA119.008729 (2019).

96. Hernandez, J. D. & Baum, L. G. Ah, sweet mystery of death! Galectins and control of cell fate. Glycobiology 12, 127R–136R, doi:10.1093/glycob/cwf081 (2002).

97. Garner, O. B. & Baum, L. G. Galectin-glycan lattices regulate cell-surface glycoprotein organization and signalling. Biochem Soc Trans 36, 1472–1477, doi:10.1042/BST0361472 (2008).

98. Rabinovich, G. A. & Croci, D. O. Regulatory circuits mediated by lectin-glycan interactions in autoimmunity and cancer. Immunity 36, 322–335, doi:10.1016/j.immuni.2012.03.004 (2012).

99. Baum, L. G., Garner, O. B., Schaefer, K. & Lee, B. Microbe-Host Interactions are Positively and Negatively Regulated by Galectin-Glycan Interactions. Front Immunol 5, 284, doi:10.3389/fimmu.2014.00284 (2014).

100. Colomb, F., Giron, L. B., Trbojevic-Akmacic, I., Lauc, G. & Abdel-Mohsen, M. Breaking the Glyco-Code of HIV Persistence and Immunopathogenesis. Curr HIV/AIDS Rep 16, 151–168, doi:10.1007/s11904-019-00433-w (2019).

101. Bradshaw, C. S. et al. High recurrence rates of bacterial vaginosis over the course of 12 months after oral metronidazole therapy and factors associated with recurrence. J Infect Dis 193, 1478–1486, doi:10.1086/503780 (2006).

102. Secura, G. M., Allsworth, J. E., Madden, T., Mullersman, J. L. & Peipert, J. F. The Contraceptive CHOICE Project: reducing barriers to long-acting reversible contraception. Am J Obstet Gynecol 203, 115 e111–117, doi:10.1016/j.ajog.2010.04.017 (2010).

103. Mark, S. S. & A., D. Analysis of Carbohydrates/Glycoproteins by Mass Spectrometry. In Cell Biology (Third Edition). Editor: Celis, J.E. Academic Press; 415–425, doi.org/10.1016/B978-012164730-8/50238-0 (2006).

104. Hara, S., Takemori, Y., Yamaguchi, M., Nakamura, M. & Ohkura, Y. Fluorometric high-performance liquid chromatography of *N*-acetyl- and *N*-glycolylneuraminic acids and its application to their microdetermination in human and animal sera, glycoproteins, and glycolipids. Anal Biochem 164, 138–145, doi:10.1016/0003-2697(87)90377-0 (1987).

105. Bigge, J. C. et al. Nonselective and Efficient Fluorescent Labeling of Glycans Using 2-Amino Benzamide and Anthranilic Acid. Analytical Biochemistry 230, 229–238, doi:DOI 10.1006/abio.1995.1468 (1995).

106. Dell, A. et al. Mass spectrometry of carbohydrate-containing biopolymers. Methods Enzymol 230, 108–132, doi:10.1016/0076-6879(94)30010-0 (1994).

107. Ceroni, A., Dell, A. & Haslam, S. M. The GlycanBuilder: a fast, intuitive and flexible software tool for building and displaying glycan structures. Source Code Biol Med 2, 3, doi:10.1186/1751-0473-2-3 (2007).

108. Ceroni, A. et al. GlycoWorkbench: a tool for the computer-assisted annotation of mass spectra of glycans. J Proteome Res 7, 1650–1659, doi:10.1021/pr7008252 (2008).

109. Choudhury, B., Carlson, R. W. & Goldberg, J. B. Characterization of the lipopolysaccharide from a wbjE mutant of the serogroup O11 Pseudomonas aeruginosa strain, PA103. Carbohydr Res 343, 238–248, doi:10.1016/j.carres.2007.11.003 (2008).

110. Peter Albersheim, D. J. N., Patricia D. English, Arthur Karr. A method for the analysis of sugars in plant cell-wall polysaccharides by gas-liquid chromatography. Carbohydrate Research 5, 340–345 (1967).

